# Extracting adverse event nature, severity, timelines and resulting interventions from clinical notes of patients receiving CAR-T therapy using large language models

**DOI:** 10.64898/2026.04.28.26351782

**Authors:** Jordan Guillot, Brenda Miao, Arvind Suresh, Madhumita Sushil, Christopher YK Williams, Rohit Vashisht, Tomiko T. Oskotsky, Marina Sirota, Atul J. Butte

## Abstract

Chimeric Antigen Receptor T-cell (CAR-T) therapy, where genetically engineered patient T cells target tumor antigens, has transformed care for hematologic malignancies but requires careful tracking of adverse events (AEs) often documented only in unstructured EHR notes.

We evaluated a Large Language Model (LLM)–based approach in UCSF’s secure environment to extract AEs, dates, grades, and interventions within 30 days post-infusion for six commercial CAR-T products (2012–2023), benchmarking against two evaluators. Using GPT-4-0314 in a zero-shot setting with four prompts (prespecified AEs, non-prespecified AEs, CRS, ICANS), we compared outputs against dual annotations on a random sample of 50 notes using accuracy, precision, recall, F1, and Cohen’s kappa. From 4,762 progress notes for 293 patients (median age 65.6), CRS occurred in 80.2% (median onset 4 days); neutropenia 70.0% (16 days); neutropenic fever 64.8% (4 days); ICANS in 34.8%. Interventions included tocilizumab and corticosteroids. Grades were frequently undocumented (CRS 62.3%, ICANS 56.1%); documented cases were mainly CRS grade 1 (59.4%) and ICANS grade 2 (28.0%). Performance was high on CRS and ICANS grading (accuracy of 0.97 and 0.91, respectively). Moderate performances were assessed for prespecified AE extraction (accuracies 0.62–0.76), and non-prespecified AEs (accuracies 0.76–0.84). Inter-rater reliability was strong to near-perfect for CRS/ICANS presence and grade (kappa 0.86–0.96), moderate for dates and interventions, and weaker for broader AE attributes.

LLM-derived insights can augment AE monitoring and real-world evidence generation by unlocking unstructured clinical detail and characteristic timelines after CAR T. However, performance varied for broader AE attributes, warranting cautious use. Performance was highest for detecting the presence and grade of CRS and ICANS, with strong to near-perfect inter-rater reliability. While cautious use of LLMs for broad AE extraction is warranted due to the variable performance observed in this study, these results support integrating high-performing CRS/ICANS extraction into EHR workflows.

**Author summary:** Chimeric Antigen Receptor T-cell (CAR-T) therapy has transformed care for blood cancer but requires careful tracking of adverse events (AEs). We asked whether a large language model could read routine clinical notes and extract AEs after CAR T-cell therapy. We analyzed de-identified notes from the first month after infusion. The model identified when two key side effects occurred—cytokine release syndrome (a whole-body inflammatory reaction) and neurotoxicity (brain and nerve symptoms)—and how severe they were, with accuracy similar to human reviewers. It also captured when side effects started and what treatments were given, though performance was more variable for the wider range of side effects beyond these two. In our data, these reactions often arose within the first week; blood count problems and infections were also common. Because many notes did not state severity explicitly, the model sometimes could not assign a grade. Our findings suggest that language models can help unlock important details hidden in clinical notes and could be incorporated into electronic records to support faster, more reliable side-effect monitoring and research. We recommend careful, supervised use and continued validation, especially for broader side-effect categories.

## Introduction

Chimeric Antigen Receptor T-cell (CAR-T) therapy represents a significant advancement in the field of oncology, offering a treatment modality that can induce high response rates and durable remissions in refractory hematologic cancers, with active innovation aimed at expanding targets, improving safety, and overcoming barriers in solid tumors (1–4). This therapeutic approach involves the genetic modification of a patient’s T-cells to express a CAR that targets specific antigens on tumor cells, thereby enhancing the immune system’s ability to recognize and destroy cancer cells (5). In 2023, six CAR T-cell therapies were approved by the Food and Drug Administration (FDA) for the treatment of hematologic malignancies, including lymphomas, some forms of leukemia, and multiple myeloma (6). These FDA-approved CAR T-cell therapies target one of two antigens on B cells, CD19 or BCMA. Despite their considerable therapeutic potential, CAR-T cell therapy are associated with a high rate of adverse events (AEs) (1–4), including Cytokine Release Syndrome (CRS)(7–9) and Immune Effector Cell-Associated Neurotoxicity Syndrome (ICANS) (10). which can occur frequently among patients receiving these therapies.

CRS is a severe and potentially life-threatening AE characterized by an excessive immune response triggered by the activated CAR-T cells releasing large amounts of cytokines into the bloodstream (11). Symptoms of CRS can range from constitutional symptoms to severe inflammatory responses, which can lead to multi-organ dysfunction or even death if not managed promptly and effectively (12). The severity of CRS is often correlated with the disease burden and treatment response, which complicates the clinical management of affected patients (13). In addition to CRS, other AEs can be detrimental to patients, including ICANS - a neurologic toxicity of CAR-T and other immune effector therapies marked by encephalopathy, confusion, aphasia, tremor, seizures, and, rarely, cerebral edema - that is thought to arise from cytokine-driven endothelial activation and blood–brain barrier disruption. (8,10,14). Given the serious nature of CRS, ICANS, and other associated AEs, continuous and meticulous monitoring is imperative for the successful implementation of CAR-T therapy. Monitoring is essential for early detection of AEs and optimizing treatment protocols to reduce patient risks and ensure CAR-T therapy’s benefits (15).

Electronic health records (EHRs) provide a rich data source for monitoring outcomes in patients receiving CAR-T therapy; however, much of the clinically relevant information required is difficult to capture from structured data alone. Structured EHR data (diagnosis codes, medication administrations, lab values, procedure codes) are often not sufficiently accurate or detailed. As a result, they rarely capture adverse events with sufficient precision: the timeline, how symptoms change, what interventions occur, and the differences among AE grades (for example, CRS and ICANS). Prior studies have shown that relying exclusively on structured data can lead to underreporting or misclassification of adverse events in oncology cohorts, as coding practices vary across institutions and key clinical details are frequently documented only in clinical notes (16,17). In contrast, unstructured data—including provider progress notes, nursing documentation, pathology reports, and consult notes—contain the granular qualitative information necessary to identify event onset, progression, and severity. Extracting such information at scale has historically required intensive manual chart review, which is labor-intensive and not feasible for large real-world datasets (18).

The advent of large language models (LLMs), such as the GPT (Generative Pre-trained Transformer) series developed by OpenAI, allows for innovative application of artificial intelligence in various fields, including healthcare (19). LLMs are a type of artificial intelligence that utilize deep learning techniques to understand and generate human-like text based on the training received from a vast corpus of text data (20). These models are not only capable of analyzing context and generating text but also of performing tasks like translation, summarization, and, importantly for healthcare science, information extraction (21). LLMs can be used to automate and enhance various data-intensive tasks that traditionally require extensive human labor, such as extracting patient information from unstructured clinical notes or assisting in real-time patient monitoring (22–24). Leveraging natural language processing and large language models enables automated extraction of clinically relevant features from unstructured text, bridging the gap between codified data and nuanced clinical phenomena, and ultimately supporting more accurate AE monitoring in CAR-T populations (25–27).

The aim of this study was to evaluate the capability of a large language model to extract information about AEs, including their dates and associated clinical interventions, and to identify CRS and ICANS grades from the unstructured text of clinical progress notes following CAR-T therapy.

## Methods

In this cross-sectional study, we identified all patients visiting the University of California, San Francisco (UCSF) hospital between January 1, 2012 and October 21, 2023 and receiving any of the six following CAR-T cell therapies: axicabtagene ciloleucel, idecabtagene vicleucel, ciltacabtagene autoleucel, lisocabtagene maraleucel, tisagenlecleucel, and brexucabtagene autoleucel (Figure 1). From reference papers and expert opinions we identified a list of 25 adverse events that are recognized as CAR-T adverse events (6–8,14). We defined these as the prespecified adverse events, which included: “Cytokine Release Syndrome (CRS)”, “Immune Effector Cell-Associated Neurotoxicity Syndrome (ICANS)”, “Hypogammaglobulinemia”, “Neutropenic Fever”, “Macrophage Activation Syndrome (MAS)”, “Hemophagocytic Lymphohistiocytosis (HLH)”, “Tumor Lysis Syndrome”, “Movement disorder”, “Cognitive impairment”, “Personality changes”, “B-cell aplasia”, “thrombocytopenia”, “anemia”, “neutropenia”, “leukopenia”, “CD4 lymphopenia”, “Cranial Nerve Palsy”, “Bacterial infection”, “Viral Infection”, “Zoster Infection”, “Fungal infection”, “Aspergillus infection”, “Pneumocystis Jirovecii Pneumonia infection”, “Secondary myeloid malignancies”, “Graft-versus-host disease (GVHD)”.

**Figure 1.**
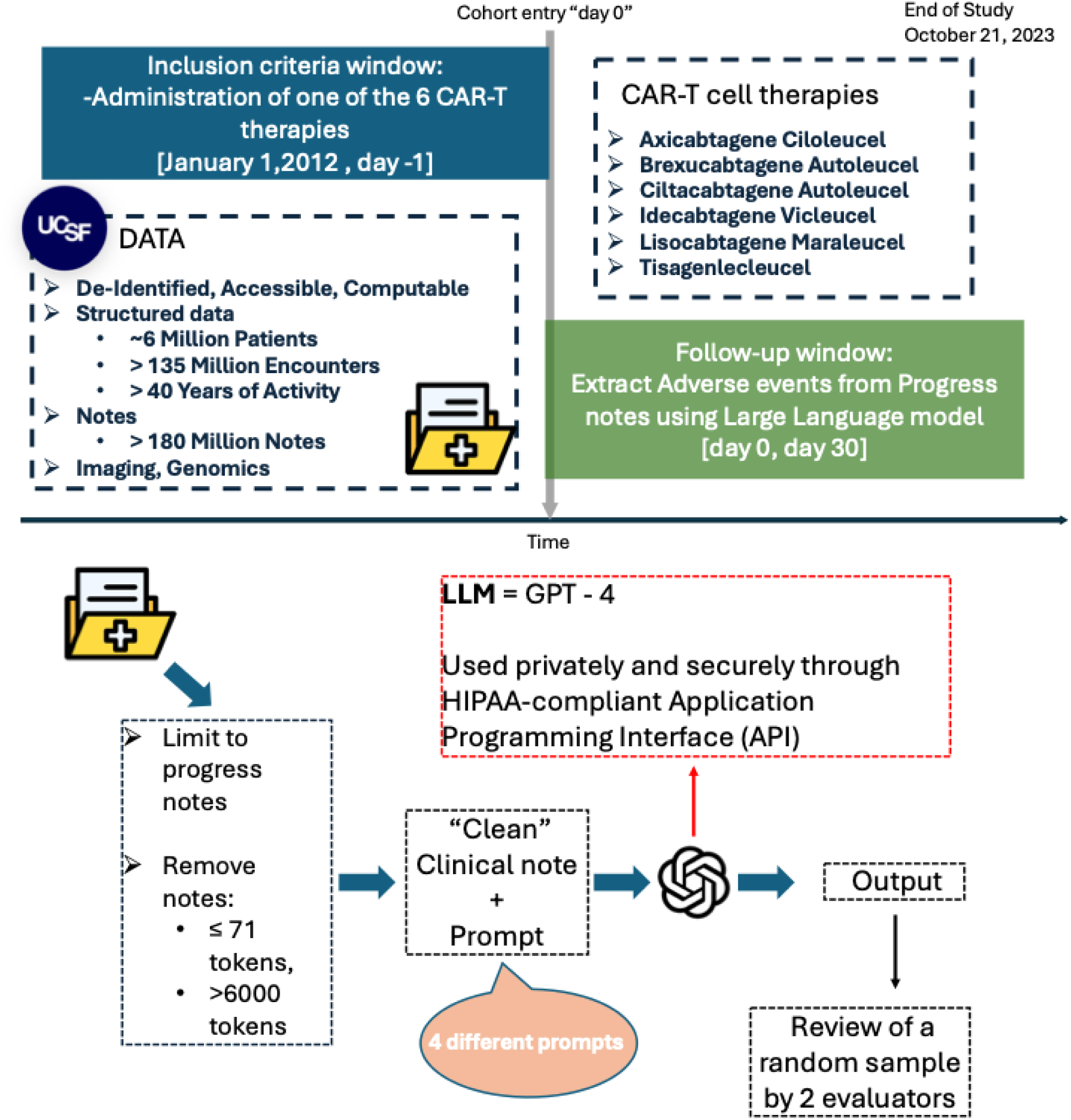
Data sources describing UCSF patient, encounter, and note counts; cohort selection based on CAR-T therapy administration with follow-up for adverse event extraction; and study workflow including note filtering, GPT-4 prompting, and random-sample annotator review Data sources and counts. The figure summarizes the electronic health record sources used at University of California, San Francisco (UCSF), including the total number of unique patients, encounters, and clinical notes available for analysis. Patient counts refer to unique individuals; encounter counts refer to distinct inpatient or outpatient visits; note counts refer to individual clinical documents (progress notes, discharge summaries, procedure notes, and related documentation). Cohort selection for chimeric antigen receptor T cell therapy recipients. The figure shows identification of the study cohort by selecting patients who received chimeric antigen receptor T cell (CAR T) therapy. The index date is defined as the date of CAR T administration. The follow up window for adverse event assessment begins at the index date and extends for 30 days after infusion, during which notes are evaluated for adverse events. Study workflow for adverse event extraction. The panel outlines the analytic workflow applied to clinical notes from the final cohort: (1) note filtering by time window relative to CAR T infusion and by note type (progress notes only); (2) large language model (LLM) prompting using Generative Pretrained Transformer 4 (GPT 4) to extract adverse event; and (3) random sample evaluator review to assess model performance. Abbreviations: UCSF, University of California, San Francisco; CAR T, chimeric antigen receptor T cell; AE, adverse event; EHR, electronic health record; LLM, large language model; GPT 4, Generative Pretrained Transformer 4.

We retrieved all clinical notes from this cohort, tokenized the text using tiktoken, and excluded notes with fewer than 71 tokens (removed bottom 25% notes, eliminating short notes with little clinical relevance) or more than 6000 tokens (corresponding to the maximum context window of the LLM used in this study). We then extracted all progress note that occurred in the 30 days following any CAR-T therapy infusion. If a patient received multiple CAR-T infusions at UCSF, each infusion was counted as a separate, independent episode. These clinical notes have been previously deidentified, externally certified, and made available for research as previously described (28). The UCSF Institutional Review Board determined that this use of deidentified structured and clinical text data in the UCSF Information Commons is not considered human participant research and that the study was therefore exempt from further approval and the need for informed consent (Figure 1).

Accessing the application programming interface (API) via the Health Insurance Portability and Accountability Act (HIPAA)–compliant UCSF Secure Azure OpenAI environment, we utilized the GPT-4 model (OpenAI model = “gpt-4-0314”; role = “user”; temperature = 0; all other settings at default values). We used four distinct prompts in a zero-shot manner (Table S1), i.e. providing no additional training context, examples or additional guidelines to the model. The first prompt asked the model to return prespecified AEs expected after any CAR-T infusion along with their dates and any associated clinical interventions documented within the clinical notes. The second prompt asked the model to detect the presence of any AEs related to CAR-T that are not among the prespecified AEs along with their dates and any clinical interventions. Third, we prompted the LLM to identify the presence, date, and grade of CRS along with any associated clinical interventions. Lastly, we prompted the LLM to identify the presence, date, and grade of ICANS, and any associated clinical interventions.

A sample of 50 notes from 47 different patients was randomly selected. LLM outputs were manually reviewed and evaluated as True or False for AEs, grades (for the 3rd and 4th prompts), dates of occurrence, and associated clinical interventions by Jordan Guillot (JG): a PharmD, PhD, with 5 years of postgraduate training in pharmacoepidemiology research and Arvind Suresh (AS): an MD, an oncologist with 3 years of postgraduate general medical training, to compare the performance between the LLM models and human annotations. We evaluated performance separately for each element: AE presence, grade, AE date, and clinical intervention. For each element, a true positive was counted when the system correctly extracted the element as documented in the note. A false positive was counted when the system extracted an element that was not documented or extracted an incorrect element. A true negative was counted when the element was not documented in the note and the LLM made no extraction. A false negative was counted when the element was documented but the LLM failed to extract it. Performance was evaluated using accuracy, precision, recall, and F1 score, computed using their standard definitions (29). The average performance was calculated by macro-averaging the results from the two evaluators. Inter-rater reliability (IRR) between the two reviewers was assessed by Cohen’s kappa (30).

## Results

A total of 293 patients receiving CAR-T cell therapy at UCSF were included in the study. Among them, 97 (33.1%) patients received Axicabtagene Ciloleucel, 64 (21.8%) received Ciltacabtagene autoleucel, 43 (14.7%) received Tisagenlecleucel, 37 (12.6%) received Lisocabtagene maraleucel, 36 (12.3%) received Idecabtagene vicleucel and 16 (5.4%) received Brexucabtagene autoleucel. Median age was 65.6 years [IQR: 54.8 – 71.7]. There were 176 (60.0%) male and 117 (40.0%) female. Among all patients 170 (58.0%) were White, 69 (23.5%) Hispanic, 34 (11.6%) Asian, 11 (3.8%) Black and ≤ 10 (≤3.4%) other than these race/ethnicity categories. Among these patients, the most common disease codes in UCSF EHR that matched the approved indications for CAR-T therapy were Diffuse Large B-Cell Lymphoma (DLBCL), coded for 150 patients (51.2%); multiple myeloma, for 131 patients (44.7%); and Acute Lymphoblastic Leukemia (ALL), for 36 patients (12.3%). Among included patients, 25,974 clinical notes documented within 30 days after CAR-T administration were identified. Specifically, after filtering by note length (i.e., retaining only notes with more than 71 tokens or fewer than 6000 tokens), 18,808 notes remained, of which 4,762 (25.3%) were progress notes retained for analysis. (Table 1). The other types of notes not retained for the analysis were 2503 (13.3%) imaging notes, 2212 (11.8%) consults, 1986 (10.6%) plans of care, 1475 (7.8%) registered nurse notes, 1359 (7.2%) interdisciplinary notes, 588 (3.1%) procedure notes and 3923 (20.9%) other type of notes.

**Table 1.**
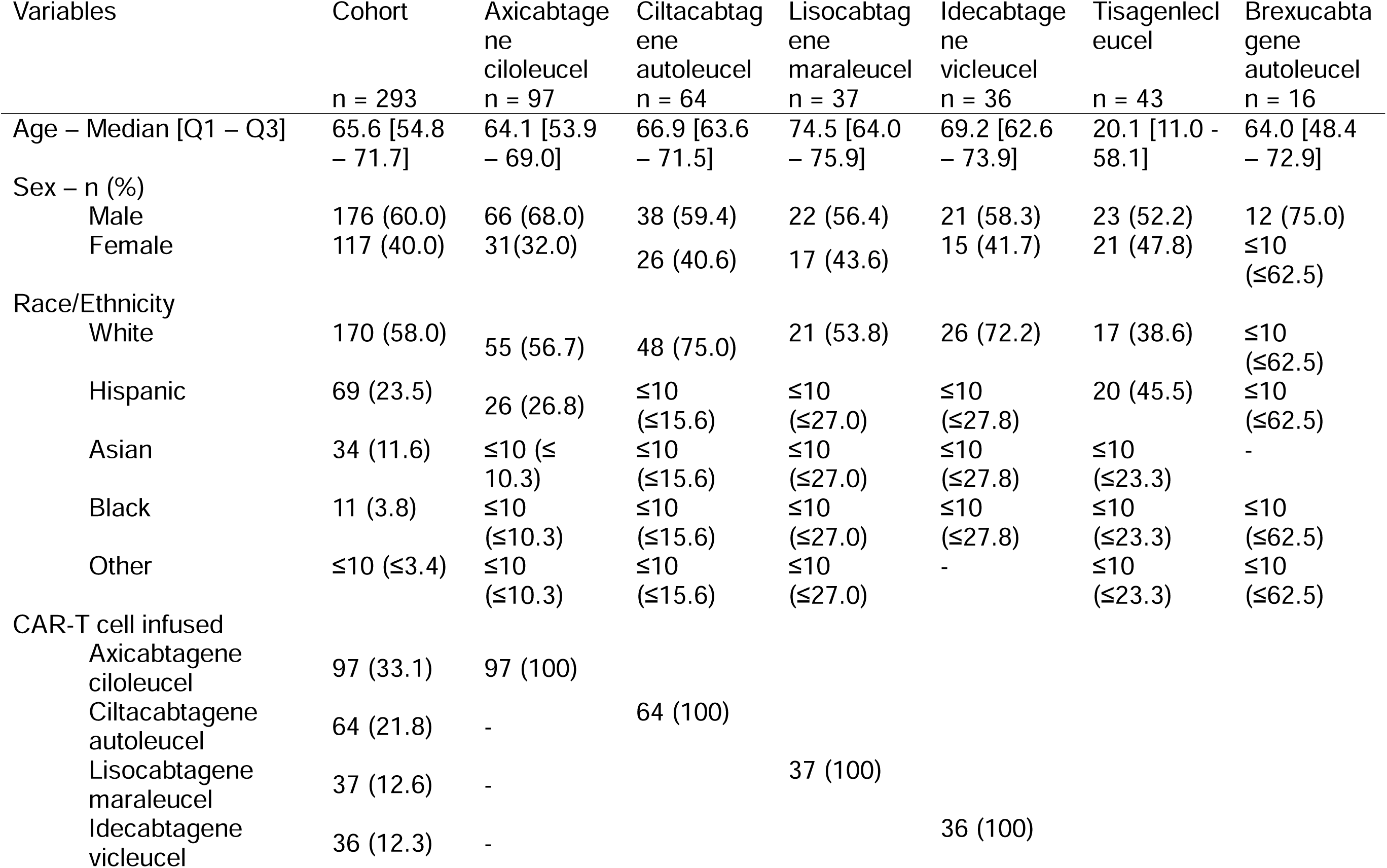
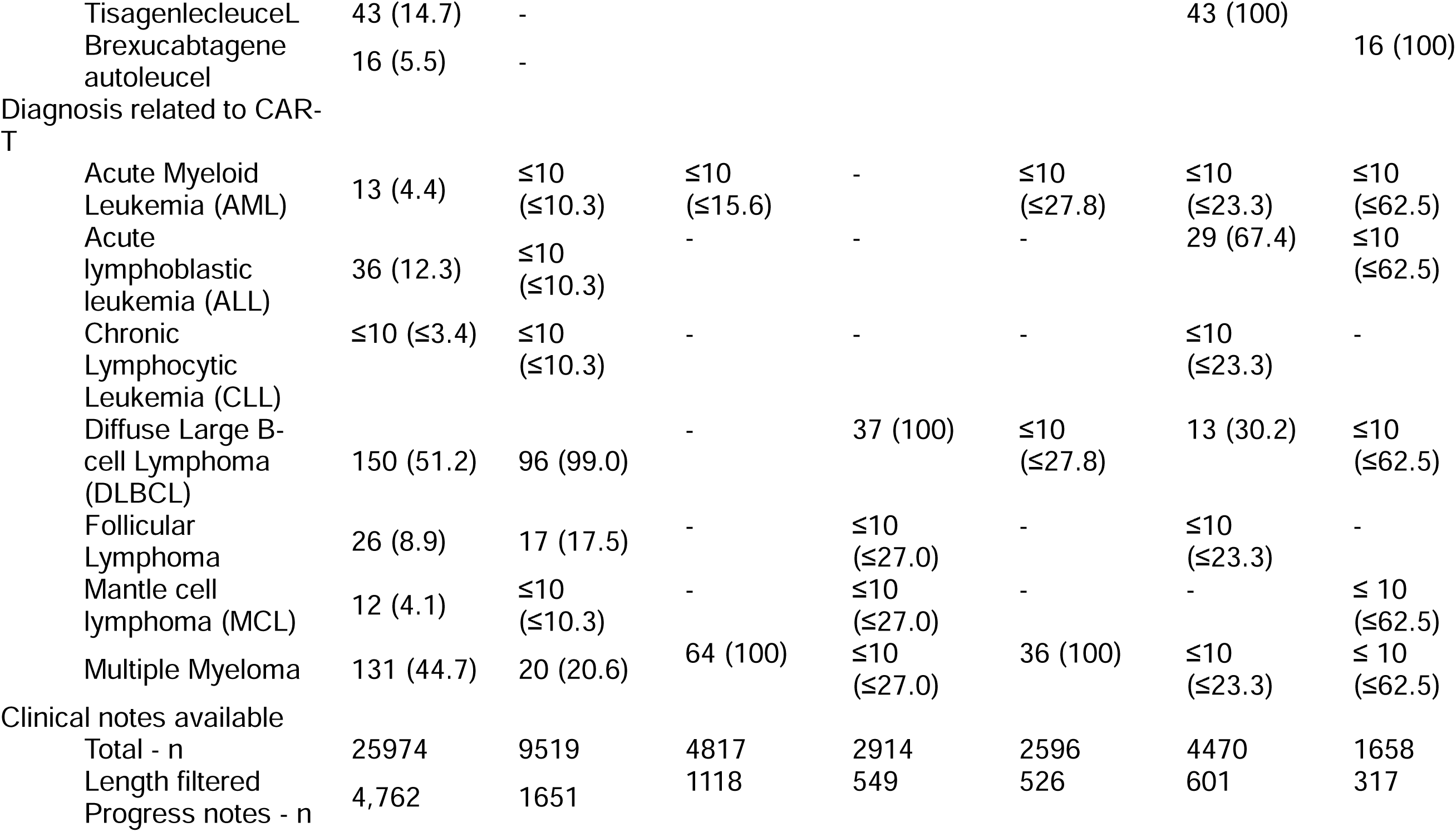
Description of the patients visiting the University of California, San Francisco (UCSF) hospital between January 1,2012 and October 21, 2023 and receiving any of the 6 following CAR-T cell therapies: axicabtagene ciloleucel, idecabtagene vicleucel, ciltacabtagene autoleucel, lisocabtagene maraleucel, tisagenlecleucel, brexucabtagene autoleucel

Performance varied across the four prompts (Figure 2, Table S2). Averaging across the two evaluators, prespecified AEs showed mean accuracies of 0.62 (nature), 0.76 (date), and 0.69 (clinical intervention), with average F1 scores ranging from 0.74 (nature) to 0.78 (date). For non-prespecified AEs, average accuracies were 0.76 (nature), 0.78 (date), and 0.84 (clinical intervention), with average F1 scores from 0.62 (date) to 0.75 (clinical intervention). For CRS-specific prompts, performance was consistently high: average accuracies ranged from 0.86 (date) to 0.97 (grade), with average F1 scores from 0.89 (date) to 0.97 (grade). For ICANS-specific prompts, average accuracies ranged from 0.69 (date) to 0.91 (grade), with average F1 scores from 0.56 (date) to 0.80 (grade).

**Figure 2.**
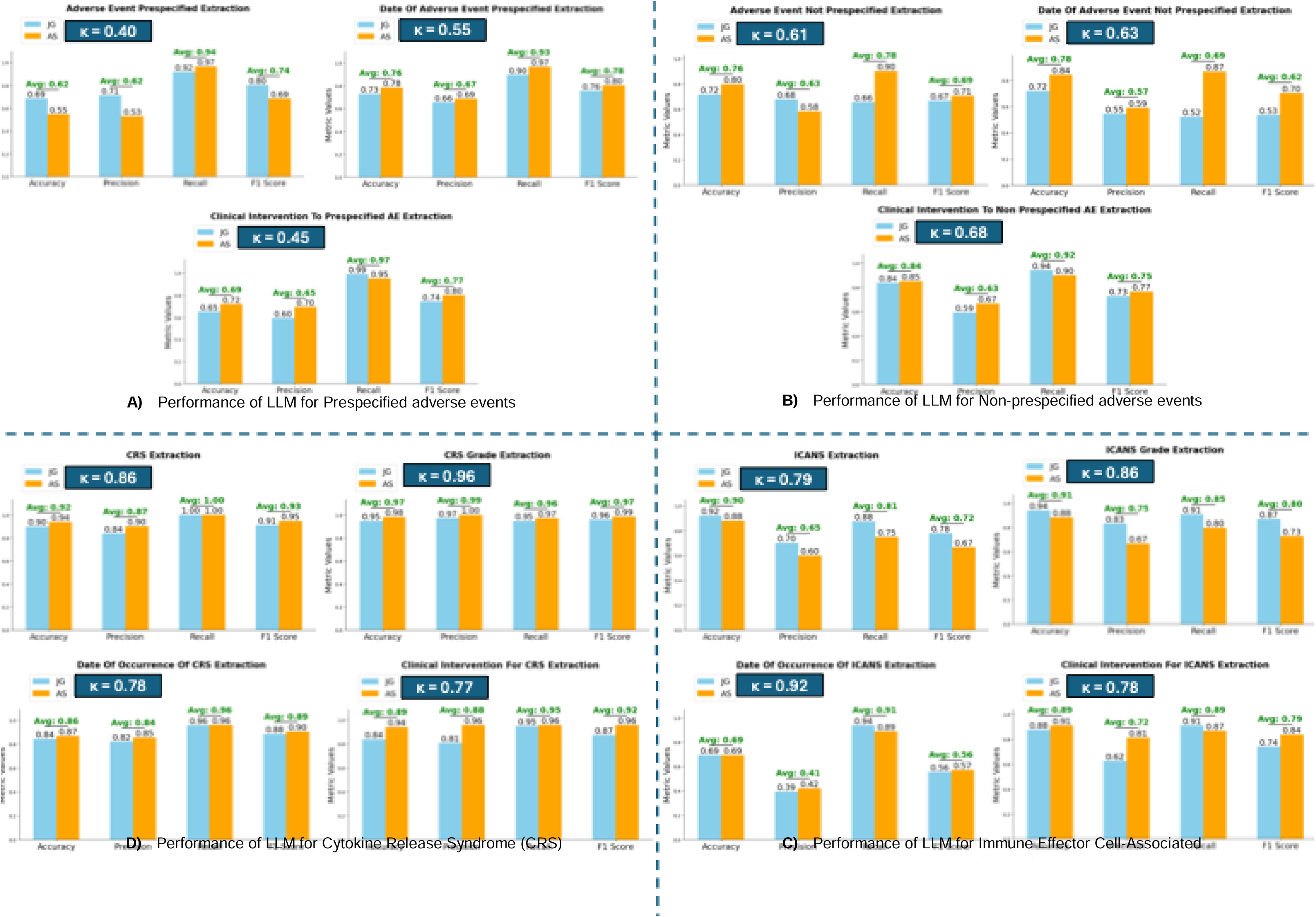
Performance of the LLM in extracting adverse event presence, grade when applicable, date, and clinical interventions from CAR-T clinical notes across four zero-shot prompts: (A) prespecified AEs, (B) non-prespecified AEs, (C) Cytokine Release Syndrome (CRS), and (D) Immune Effector Cell-Associated Neurotoxicity Syndrome (ICANS) Performance metrics include accuracy, precision, recall, and F1 score. Blue bars represent results from reviewer Jordan Guillot (JG), and orange bars represent results from reviewer Arvind Suresh (AS). Green text above each metric indicates the average value across evaluators. The blue box in each panel displays Cohen’s kappa value, representing inter-reviewer agreement. Abbreviations: adverse event, AE; cytokine release syndrome, CRS; immune effector cell-associated neurotoxicity syndrome, ICANS; F1 score, harmonic mean of precision and recall; Cohen’s kappa, measure of inter-reviewer agreement.

Inter-rater reliability (IRR), assessed using Cohen’s kappa, between evaluators varied by output category, using the bands weak (0.40–0.60), moderate (0.60–0.80), strong (0.80–0.90), and near-perfect (>0.90). For prespecified AEs, IRR was weak (nature 0.40; date 0.55; clinical intervention 0.45). For non-prespecified AEs, IRR was moderate (nature 0.61; date 0.63; clinical intervention 0.68). For CRS prompts, IRR was strong for presence (0.86), near-perfect for grade (0.96), and moderate for date (0.78) and clinical intervention (0.77). For ICANS prompts, IRR was moderate for presence (0.79) and clinical intervention (0.78), strong for grade (0.86), and near-perfect for date (0.92). (Figure 2).

The most common prespecified AEs among all patients included CRS, observed in 235 patients (80.2%), with a median onset of 4 days (IQR: 2–7); neutropenia, affecting 205 patients (70.0%), with a median onset at 16 days (IQR: 9–22); and neutropenic fever, reported in 190 patients (64.8%), with a median onset of 4 days (IQR: 2–7). Other hematologic toxicities, such as thrombocytopenia (155, 52.9%) and anemia (148, 50.5%) were also common and had median onset timelines of 19 days (IQR: 8–24) and 15 days (IQR: 6–22), respectively. Infectious complications were also prevalent, with bacterial infections documented in 84 patients (28.7%) and viral infections in 29 patients (9.9%), both occurring at a median timeline of 7–9 days post-treatment. ICANS was reported in 102 (34.8%) patients (Figure 3, Table 2). The most common information on clinical interventions for prespecified AEs extracted by the LLM included “no transfusions needed today” (reported for 48 patients, 16.4%), tocilizumab administration (46, 15.7%), and “Transfuse for Hb<7,plt<10k” (25, 8.5%) (Figure 3, Table S3).

**Figure 3.**
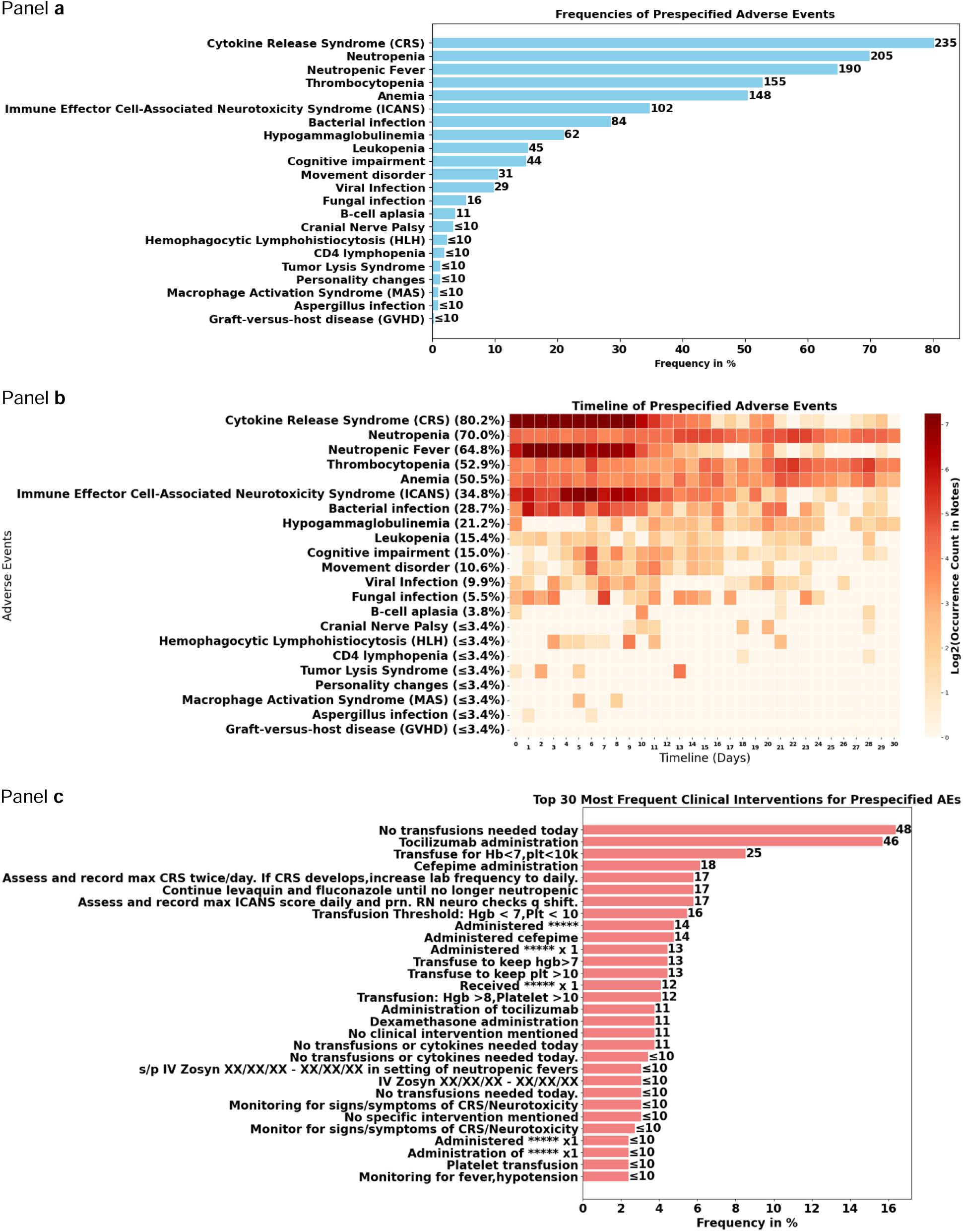
Frequencies and timelines of all identified prespecified adverse events for our cohort (n = 293 patients), with frequencies of the 30 most common resulting clinical interventions This figure presents the frequencies and timelines of prespecified adverse events (AEs) observed in a cohort of 293 patients, alongside the most common clinical interventions implemented in response. Panel a) Displays a horizontal bar chart illustrating the frequencies of prespecified adverse events as a percentage of the total cohort. The adverse events are ranked from most to least frequent. Each bar represents the percentage of patients experiencing the respective adverse event. Panel b) Shows a heatmap depicting the timeline of prespecified adverse events over the 30-day follow-up time window. The x-axis represents the timeline in days, while the y-axis lists the adverse events and their frequencies. The color intensity corresponds to the log2-transformed occurrence count in clinical notes, with darker red indicating higher frequencies. Panel c) Features a horizontal bar chart of the 30 most frequent clinical interventions for managing the prespecified adverse events. The interventions are ranked by frequency. Each bar represents the frequency patients with the intervention as a percentage of total patient number. Abbreviations used in the figure: CRS: Cytokine Release Syndrome, ICANS: Immune Effector Cell-Associated Neurotoxicity Syndrome, HLH: Hemophagocytic Lymphohistiocytosis, MAS: Macrophage Activation Syndrome, GVHD: Graft-versus-host disease

**Table 2.**
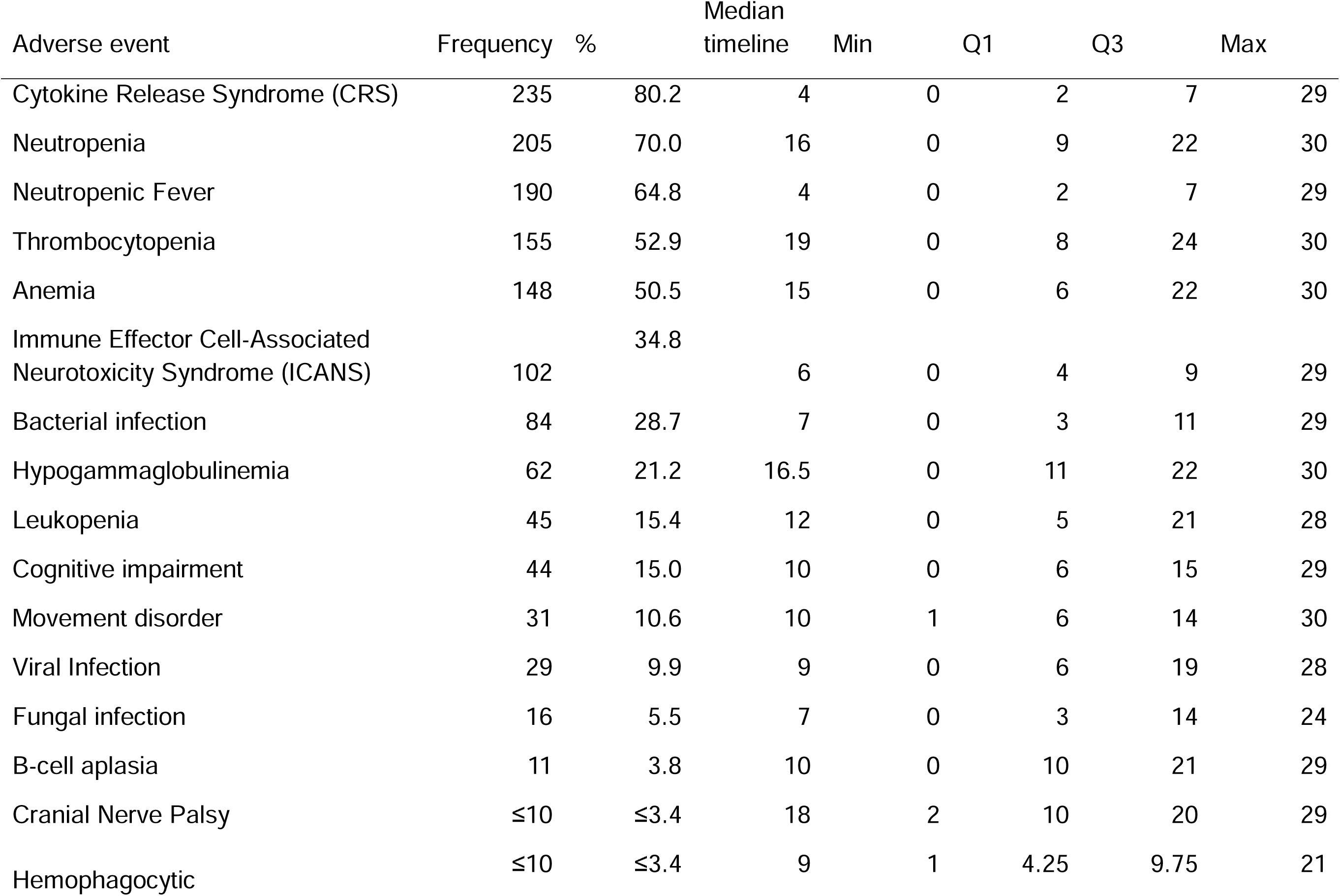
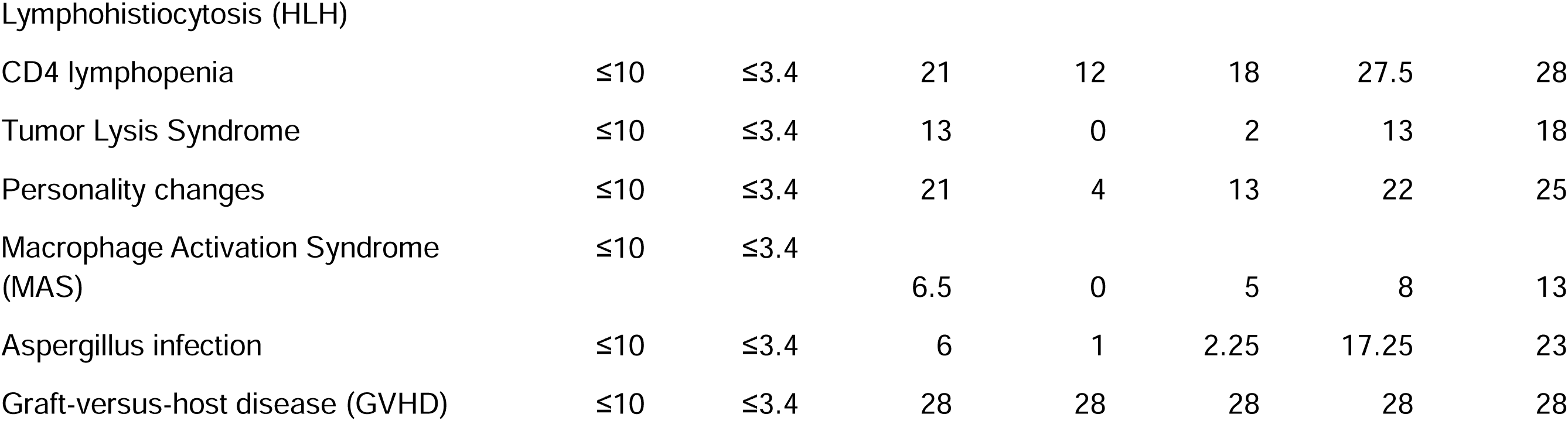
Frequencies and timeline and prespecified adverse events.

Non-prespecified AEs were less frequent than prespecified AEs and more heterogeneous in type because not classified as categories. The most prevalent included fever (4.1%), nausea (3.8%), hypotension (≤3.4%), and constipation (≤3.4%), with median onset timelines ranging from 4 to 7.5 days post-CAR-T therapy. Due to the heterogeneity of non-prespecified AEs extracted, we grouped them for descriptive purposes (Table S4). When grouped by medical domains, The most prevalent included Neurological Issues and Dizziness (20.1%), Gastrointestinal Issues (18.8%), and Cardiovascular Issues (16.7%), although they might be symptoms of prespecified AEs that would have been wrongly extracted in the prompt for non-prespecified AEs (Figure 4, Table 3). Clinical interventions for non-prespecified AEs were also heterogeneous and included continuous telemetry monitoring (CTM) (≤3.4%) and bicarbonate supplementation (≤3.4%) (Figure 4, Table S5).

**Figure 4.**
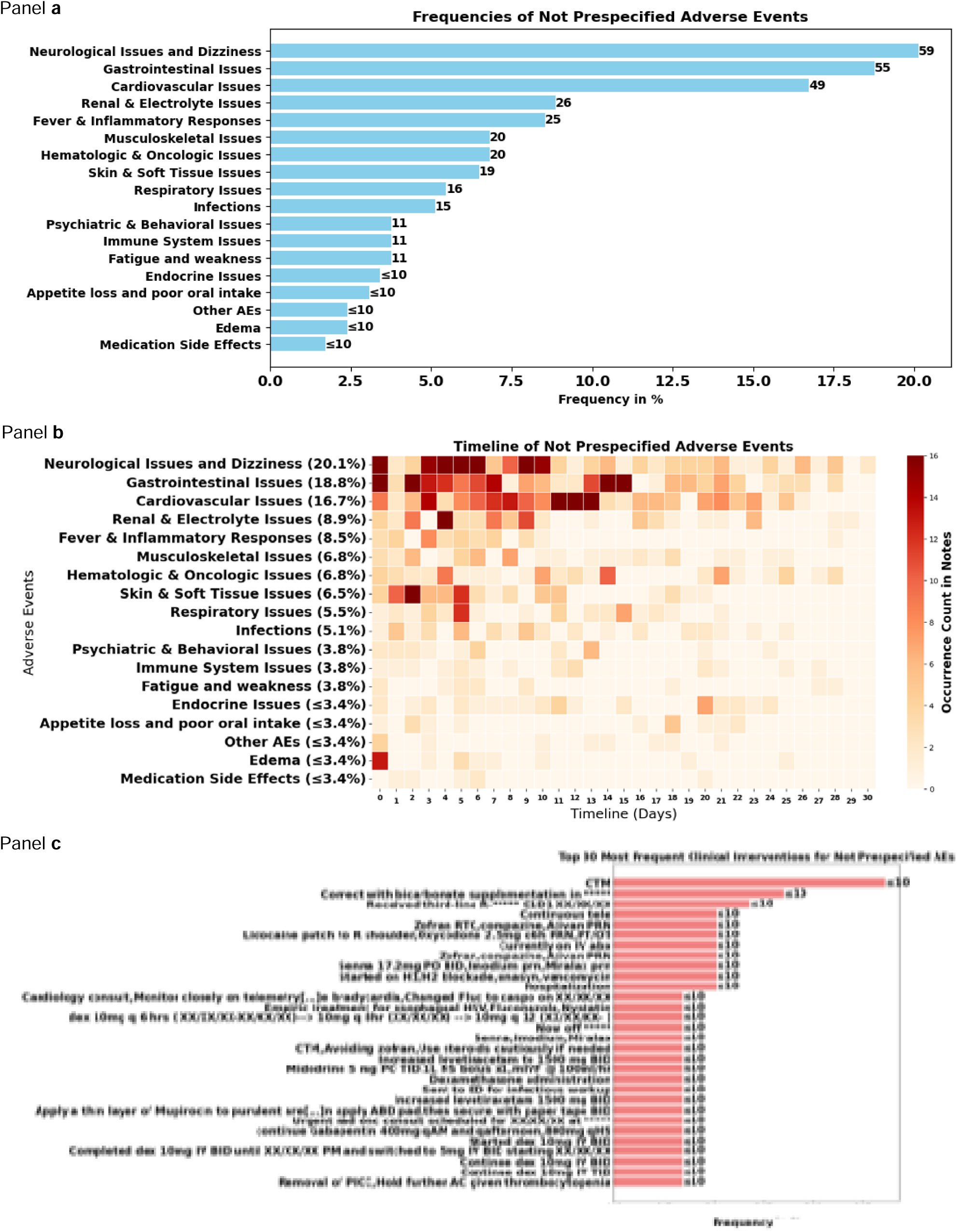
Frequencies and timelines of non prespecified adverse events for our cohort (n = 293 patients) categorized by medical categories This figure presents the frequencies and timelines of non-prespecified adverse events (AEs) observed in a cohort of 293 patients, alongside the most common clinical interventions implemented in response. Panel a) Displays a horizontal bar chart illustrating the frequencies of non-prespecified adverse events grouped by medical categories as a percentage of the total cohort. The categories are ranked from most to least frequent. Each bar represents the percentage of patients experiencing the respective adverse event. Panel b) Shows a heatmap depicting the timeline of non-prespecified adverse events grouped by medical categories over the 30-day follow-up time window. The x-axis represents the timeline in days, while the y-axis lists the adverse events grouped by medical categories and their frequencies. The color intensity corresponds to the occurrence count in clinical notes, with darker red indicating higher frequencies. Panel c) Features a horizontal bar chart of the 30 most frequent clinical interventions for managing the non-prespecified adverse events. The interventions are ranked by frequency. Each bar represents the frequency of patients with the intervention. Abbreviations used in the figure: CTM: continuous telemetry monitoring, R:Right, XX/XX/XX: dates were masked, RTC: Return to Clinic, PRN: As needed (“pro re nata”), q6h: Every 6 hours, PT/OT: Physical Therapy/Occupational Therapy, PO: By mouth (orally), BID: Twice a day, IM: Intramuscular, prn: As needed, H1/H2 blockade: Histamine-1/Histamine-2 receptor blockade, IV: Intravenous, ABD: Abdomen, q12h: Every 12 hours, q8h: Every 8 hours, qAM: Every morning, qHS: Every night at bedtime, TID: Three times a day, AC: Before meals (“ante cibum”), PICC: Peripherally Inserted Central Catheter

**Table 3.**
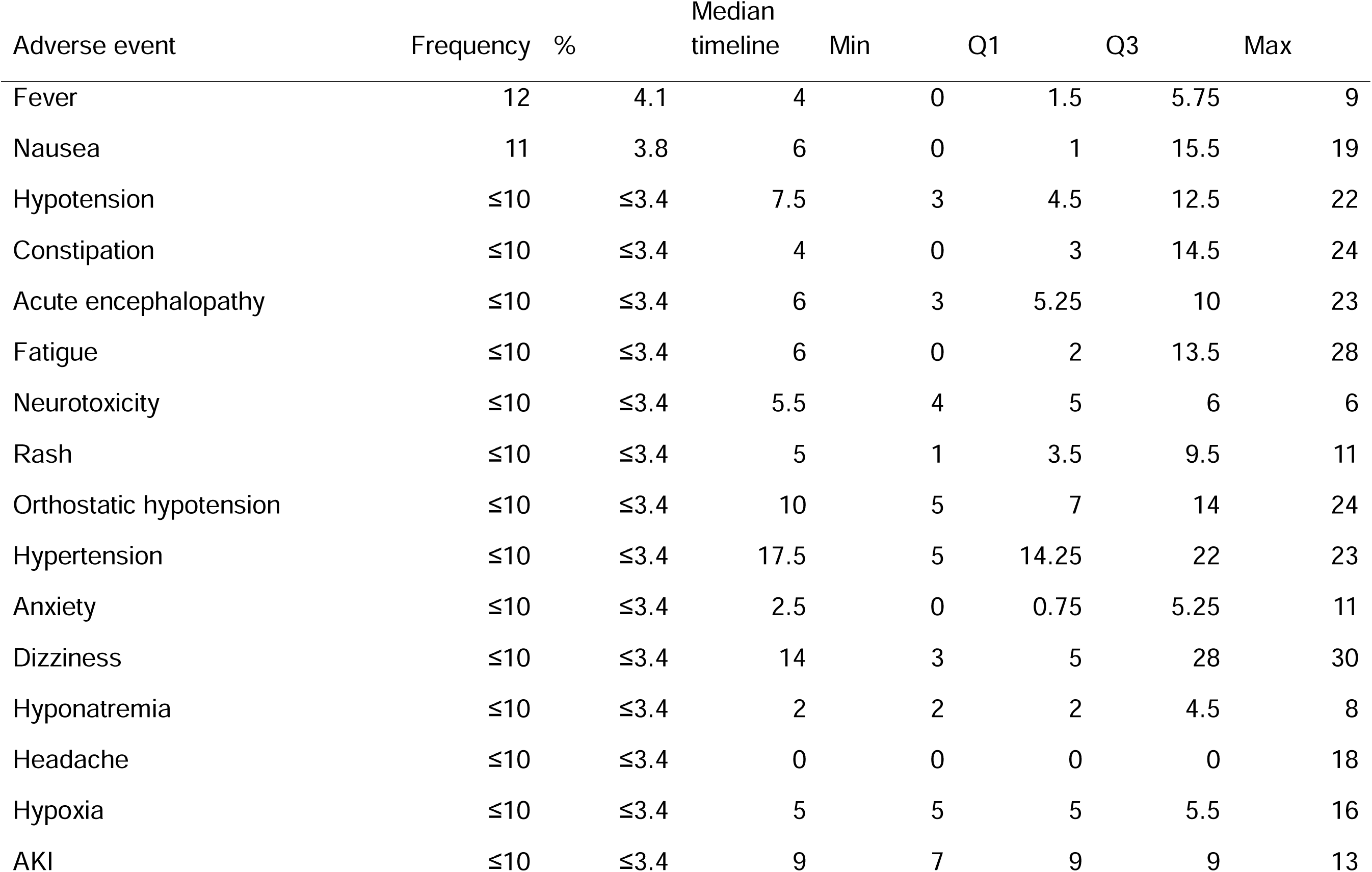
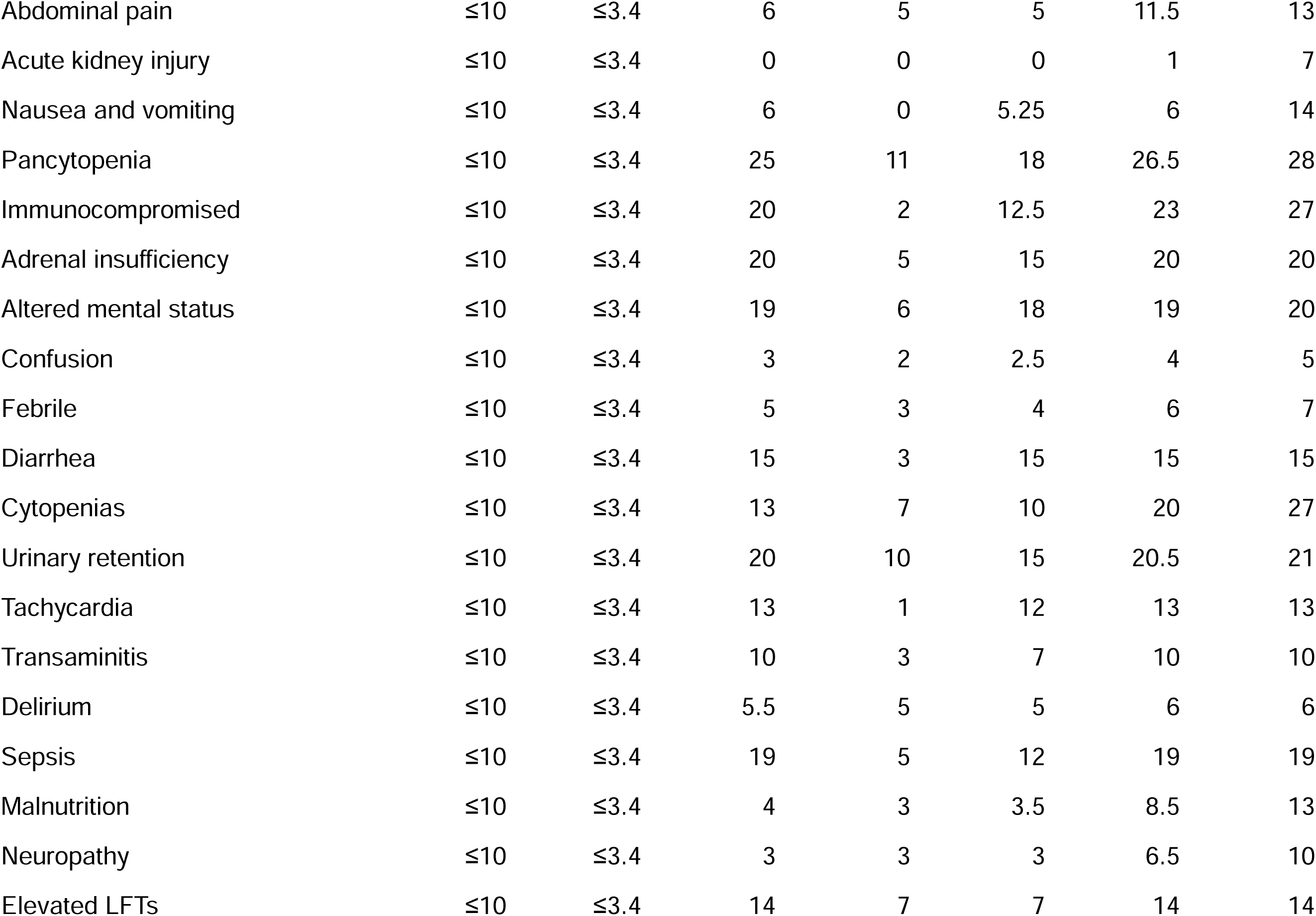
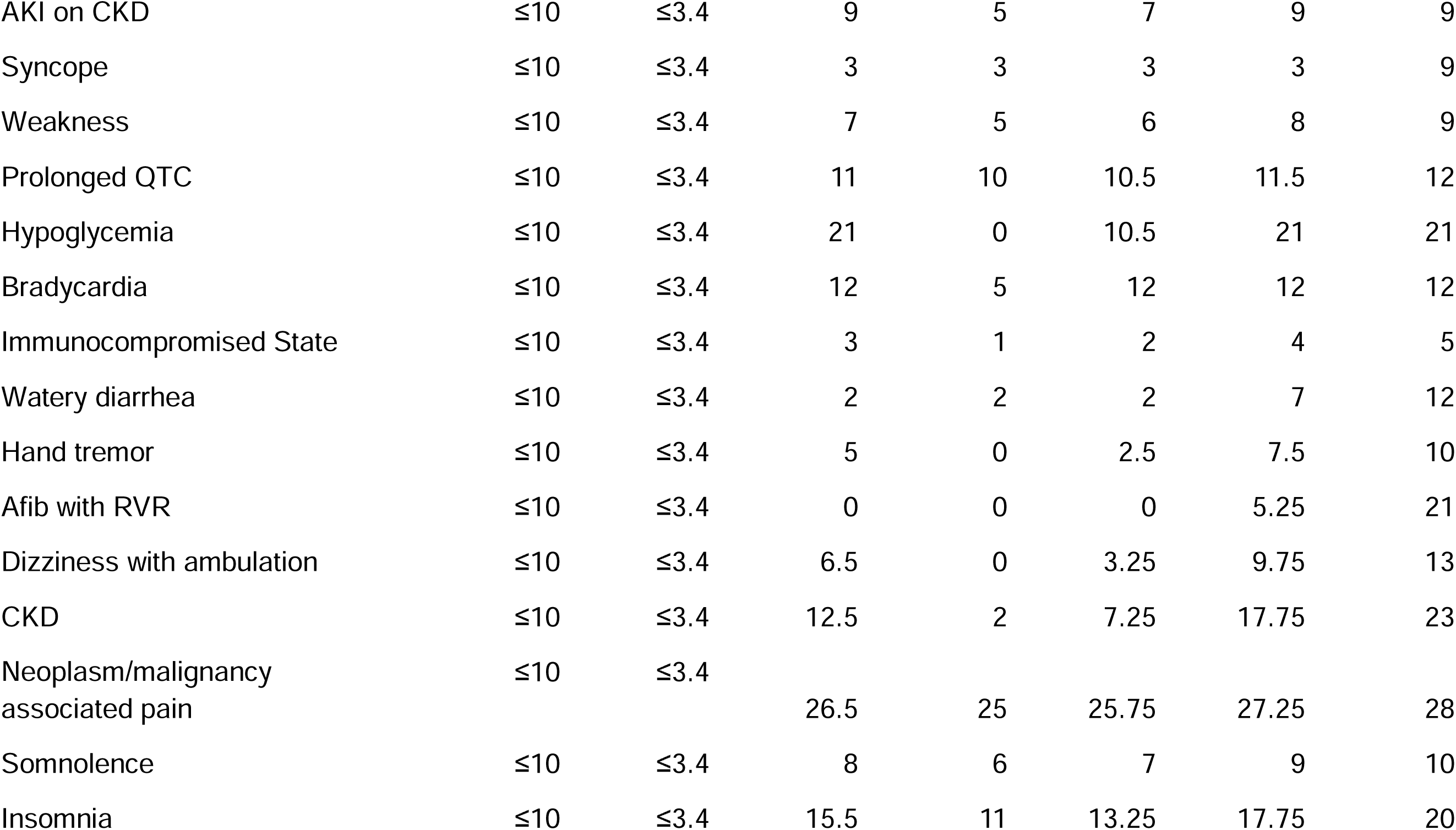
Frequencies and timeline of 50 most frequently extracted single non prespecified adverse events.

The prompt aimed at extracting the presence of CRS and its grade identified that CRS impacted 239 (81.6%) of patients. Among those, Grade 1 CRS was observed in 142 patients (59.4%), making it the most common grade reported. Grade 0 CRS occurred in 71 patients (29.7%), Grade 1–2 CRS in ≤10 patients (≤4.2%), Grade 2 CRS in 76 patients (31.8%), and Grade 2–3 CRS in ≤10 patients (≤4.2%). Grade 3 CRS in ≤10 patients (≤4.2%), and Grade 4 CRS in ≤10 patient (≤4.2%). CRS grade was identified as unknown for 148 patients (61.9%), highlighting limitations in information available in clinical notes, e.g., physician mentioning CRS without stating the grades even though they might give characteristics of the CRS. Because multiple CRS grades could be documented for the same patient across notes or time points, the grade-specific percentages are not mutually exclusive and should not be summed to 100%; rather, they reflect how frequently each grade was identified overall. (Figure 5, Table 3). The timeline for CRS onset varied across grades, with median onset days ranging from 2 to 7 days post-CAR-T therapy. Patients with Grade 1 CRS had a median onset of 4 days, whereas Grade 2 CRS had a median onset of 3 days. For Grade 3 CRS, the median timeline was 6 days, and Grade 4 CRS had a median onset of 5 days (Figure 5, Table 4). The most frequent clinical interventions were, for Grade 0 CRS, daily assessment of CRS. For Grade 1 CRS, administration of tocilizumab, for Grade 2 CRS, dexamethasone 10 mg IV Q6H, for Grade 3 CRS, administration of dexamethasone 10 mg IV Q6H, for Grade 4 CRS, the use of methylprednisone pulse therapy (Figure S1, Tables S6 – S12).

**Figure 5.**
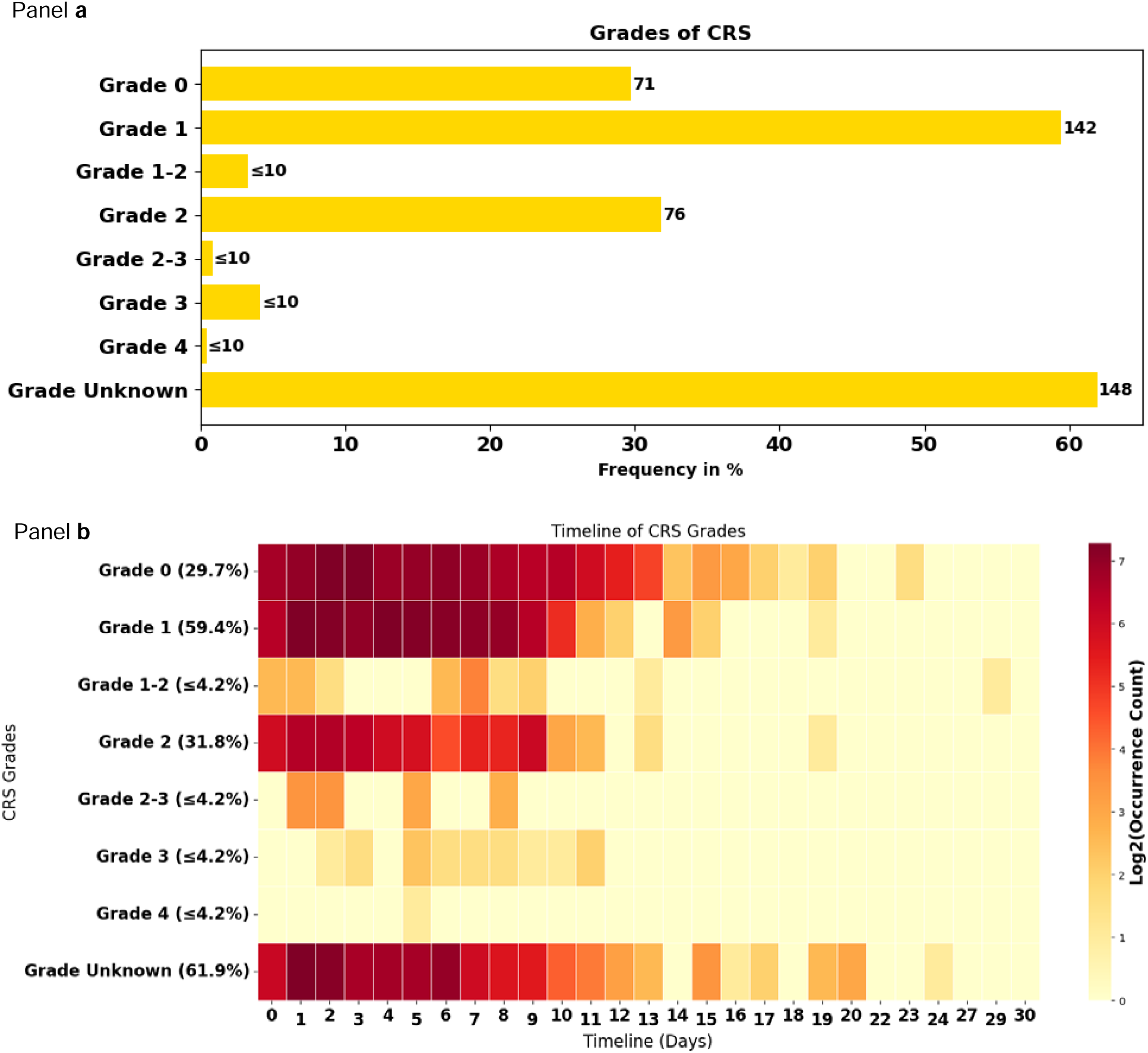
CRS grades: frequencies (bar chart) and timelines (heat map) for our patients experiencing CRS (n = 239 patients), identified using a grade-focused prompt during CRS event detection This figure presents the frequencies and timelines of occurrence of Cytokine Release Syndrome (CRS) grades in the 239 patients with CRS from our cohort of 293 patients. Panel a) Displays a horizontal bar chart illustrating the frequencies of CRS grades as a percentage of the total patients identified with CRS. The grades are ranked from least to most severe. Panel b) Shows a heatmap depicting the timeline of CRS grades over the 30-day follow-up time window. The x-axis represents the timeline in days, while the y-axis lists the CRS grades and their frequencies. The color intensity corresponds to the log2-transformed occurrence count in clinical notes, with darker red indicating higher frequencies. Abbreviations used in the figure: CRS: Cytokine Release Syndrome

**Table 4.** Frequencies and timeline of CRS Grades and ICANS Grades.

|  | Frequency | Percentage | min | q1 | median | q3 | max |
| --- | --- | --- | --- | --- | --- | --- | --- |
| CRS occurrence | 239 |  | 0 | 2 | 4 | 7 | 30 |
| CRS Grades |  |  |  |  |  |  |  |
| 0 | 71 | 29.7 | 0 | 2 | 5 | 8 | 30 |
| 1 | 142 | 59.4 | 0 | 2 | 4 | 7 | 22 |
| 1-2 | ≤10 | ≤4.2 | 0 | 1 | 7 | 7 | 29 |
| 2 | 76 | 31.8 | 0 | 2 | 3 | 7 | 20 |
| 2-3 | ≤10 | ≤4.2 | 1 | 1 | 2 | 5 | 8 |
| 3 | ≤10 | ≤4.2 | 0 | 5 | 6 | 9 | 11 |
| 4 | ≤10 | ≤4.2 | 5 | 5 | 5 | 5.5 | 6 |
| Unknown | 149 | 62.3 | 0 | 1 | 4 | 6 | 24 |
| ICANS occurrence | 132 |  | 0 | 3 | 5 | 8 | 27 |
| ICANS Grade |  |  |  |  |  |  |  |
| 0 | 55 | 41.7 | 0 | 2 | 5 | 8 | 27 |
| 1 | 33 | 25.0 | 0 | 3 | 6 | 8 | 22 |
| 1-2 | ≤10 | ≤7.6 | 6 | 6 | 6 | 6 | 6 |
| 2 | 37 | 28.0 | 0 | 4 | 6 | 8 | 23 |
| 2-3 | ≤10 | ≤7.6 | 5 | 5 | 5 | 5 | 5 |
| 3 | 12 | 9.1 | 0 | 5 | 6 | 8 | 11 |
| 3-4 | ≤10 | ≤7.6 | 6 | 6.5 | 7 | 7.5 | 8 |
| 4 | ≤10 | ≤7.6 | 6 | 6.5 | 7 | 7 | 7 |
| Unknown | 74 | 56.1 | 0 | 3 | 5 | 8 | 27 |

The prompt designed to identify ICANS grades reported that ICANS occurred in 132 (45.1%) patients. Among those, Grade 0 which correspond to absence of ICANS was the most commonly extracted information, observed in 41.7% of the 132 cases, followed by Grade 2 (28.0%) and Grade 1 (25.0%). Higher grades of ICANS, including Grade 3 (9.1%), Grade 3–4 (≤7.6%), and Grade 4 (≤7.6%), were less frequent. Grade Unknown was reported in 56.1% of cases, potentially reflecting again incomplete documentation. Because more than one ICANS grade could be recorded for an individual patient across different notes or time points, the grade-specific percentages are not mutually exclusive and should not be added to total 100%; instead, they indicate the overall frequency with which each grade was identified. (Figure 6, Table 4). ICANS median timeline was 5 to 7 days post-CAR-T therapy. (Figure 6, Table 4). The most frequent clinical intervention was seizure prophylaxis with Keppra for Grade 0 ICANS, no intervention for Grade 1, dexamethasone 10mg IV q6h for Grades 2 and 3, and Solumedrol 1 gram IV daily for 3 days for Grade 4 (Figure S2, Tables S13 – S20).

**Figure 6.**
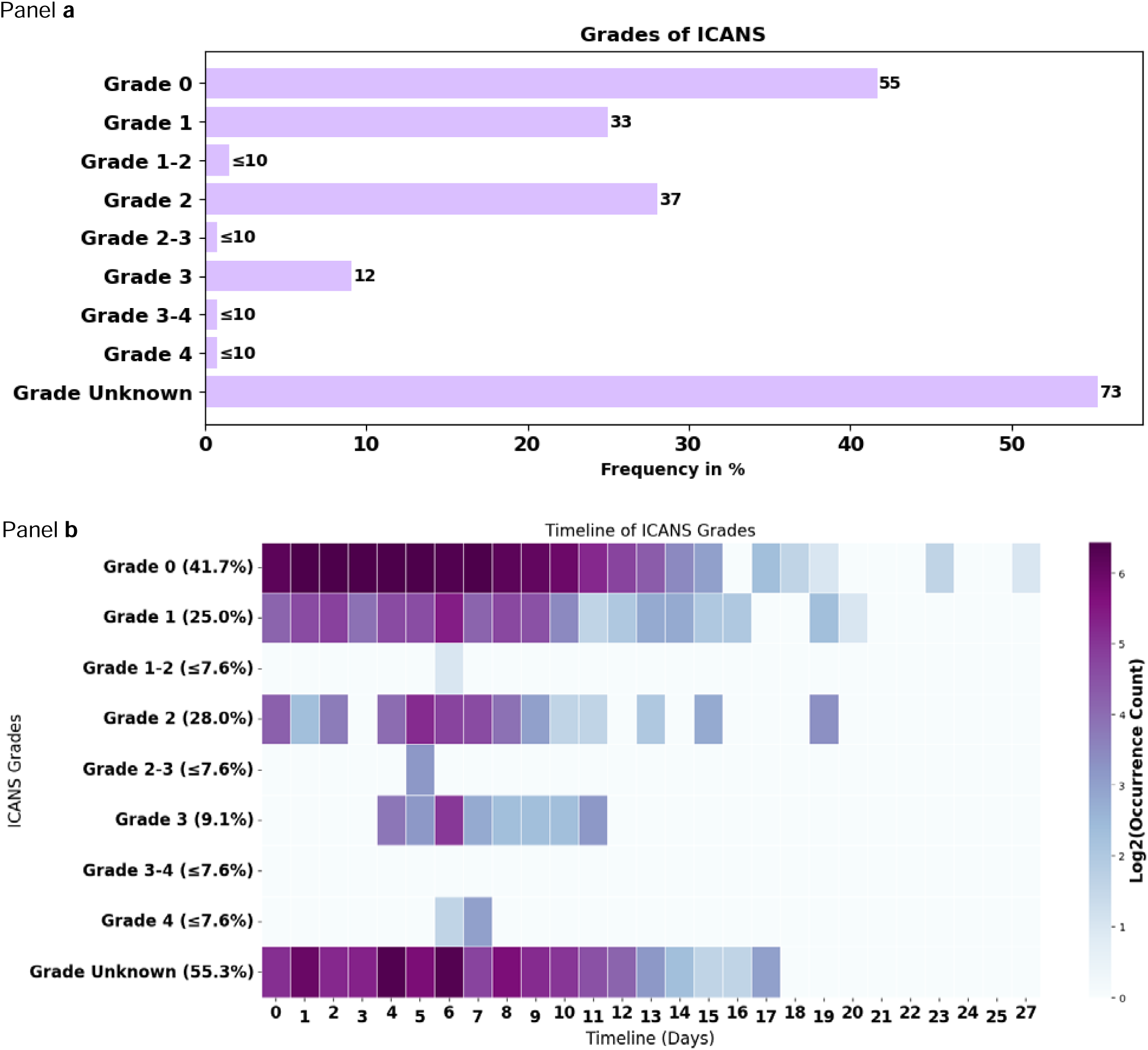
ICANS grades: frequencies (bar chart) and timelines (heat map) for our patients experiencing ICANS (n = 132 patients), identified using a grade-focused prompt during ICANS event detection This figure presents the frequencies and timelines of occurrence of Cytokine Release Syndrome (CRS) grades in the 132 patients with ICANS from our cohort of 293 patients. Panel a) Displays a horizontal bar chart illustrating the frequencies of ICANS grades as a percentage of the total patients identified with ICANS. The grades are ranked from least to most severe. Panel b) Shows a heatmap depicting the timeline of ICANS grades over the 30-day follow-up time window. The x-axis represents the timeline in days, while the y-axis lists the ICANS grades and their frequencies. The color intensity corresponds to the log2-transformed occurrence count in clinical notes, with darker purple indicating higher frequencies. Abbreviations used in the figure: ICANS: Immune Effector Cell-Associated Neurotoxicity Syndrome

## Discussion

Adverse events (AEs) after CAR-T therapy are common and often severe making timely detection and intervention essential for patient safety. However, much of the clinical detail needed to understand AE onset, severity, and management is buried in unstructured notes that are difficult to analyze at scale. Leveraging large language models (LLMs) to extract these patterns offers a promising way to better characterize AE trajectories and ultimately improve monitoring and outcomes for CAR-T recipients.

In our study, the LLM demonstrated strong performance for CRS- and ICANS-specific prompts, particularly for presence and grade. For broader prespecified AE prompts, performance was moderate and showed evaluator-dependent variability, likely reflecting the ambiguity and variability of AE documentation in unstructured clinical notes and the inherent challenges of using an LLM to identify multiple categories of adverse events. In contrast, agreement was higher for CRS and ICANS presence and grading, likely due to their more standardized clinical definitions and documentation. When using the LLM in our data, trends in AE occurrence highlighted CRS as the most frequently reported event, followed by neutropenia in prevalence, with each displaying characteristic timing patterns post-treatment. Prompts designed to extract grades provided insight into the distribution of severity, with lower grades predominating for CRS and ICANS. These results are supported by the strong accuracy and IRR; however, limitations in data completeness occasionally hindered precise grade assignment. These findings underscore the potential of LLMs to reliably capture key information on CRS and ICANS following CAR-T, while also highlighting areas where heterogeneity across other AEs and incomplete documentation may affect performance, warranting cautious use for this task.

Recent work has demonstrated the effective application of LLMs to extract clinical information from unstructured data (31–35). Moreover, emerging studies have shown good performance in the application of these techniques for adverse event detection, while emphasizing challenges such as domain-specific variability in model performance, large language models and LLM-based pipelines show strong performance for adverse drug event extraction and structured AE grades, with F1-scores between 0.62 and 0.99, outperforming traditional machine learning models and ICD-based systems (36–40). Our study aligned with these previous findings, highlighting that a commercial LLM demonstrated moderate to high performance metrics for extraction of CAR-T related AEs, similar to those reported in these prior studies that include oncology and also other domains (36–40). These results encourage physicians and researchers to leverage these tools in CAR-T monitoring, clinical trials, and real-world evidence research, while emphasizing the need for further research to improve and homogenize performance.

Across pivotal CAR-T cell therapy trials and large real-world cohorts, CRS and ICANS were frequently reported which aligned with the extraction we performed on UCSF data using LLMs. Moreover, in our study, the model captured low-frequency, non-prespecified events such as rash and constipation with good accuracy and IRR, which may serve as early warning signs of broader toxicity syndromes or signal unexpected AEs. In the pivotal ZUMA-1 trial of axicabtagene ciloleucel, the most common AEs of any grade were fever/pyrexia (85% of the patients), neutropenia (84%), and anemia (66%). CRS occurred in 93% of patients, with grade ≥ 3 events in about 13%, while neurologic AEs in 64% with 28% grade ≥ 3 (1). The JULIET trial of tisagenlecleucel reported rates of CRS (58%), anemia (48%), neutropenia (34%) and any neurologic events (12%) (3), and the TRANSCEND study of lisocabtagene maraleucel demonstrated lower frequencies of neutropenia (60%), anemia (37%), CRS (42%) and ICANS (30%) (41). Our study included several CAR-T therapies and was conducted in a real-world setting, which might explain differences to clinical trials of single drugs, although our results preserved relative proportions of the common AEs and fit what is clinically expected. Meta-analytic evidence were also supporting prevalence of these AEs, indicating pooled CRS and ICANS incidences of 70–90% and 25–30%, respectively, with pooled high-grade CRS under 15%, under 30% for all grades ICANS about 10% for high-grade ICANS (8,42). Furthermore, large multicenter registries have reported similar rates of severe CRS and ICANS, with most events occurring within the first two weeks post-infusion and resolving with tocilizumab or corticosteroids, which validate the timeline and interventions that we were able to extract (43–45). This data along with our performance analysis shows that LLMs have strong performance in identifying CRS and ICANS occurrence and their respective grades. Our study also showed the high prevalence of CRS, ICANS, neutropenia, anemia, and infections in the real-world CAR-T setting, consistent with previous clinical trials and registry-based analyses. Furthermore, we showed that these AEs typically occur early after infusion, often within the first two weeks, highlighting the critical need for close monitoring and early intervention (13,45). Severity, and current and previous care are essential considerations in the management of ongoing or evolving toxicities, and can significantly influence the trajectory and recovery from CRS, ICANS, or hematologic AE (9,10,13,43,45). Our LLM based system demonstrated high accuracy in extracting occurrence and grades CRS and ICANS from clinical notes, offering substantial potential to enhance real-time patient monitoring, clinical decision support, and pharmacovigilance monitoring. These findings support further work for integrating LLM-driven extraction tools into clinical and research settings to complement existing manual or structured use of data withing oncology practice and research.

This work has several important strengths, while also having limitations that merit consideration. Event extraction relied heavily on documentation quality, and the high proportion of “grade unknown” for CRS and ICANS highlights incomplete or inconsistent charting. This highlights variability in the way physicians document their notes and suggests that grades are not always explicitly stated but are often expected to be inferred by clinicians reading one another’s notes. This is a calling for further research into LLM interpretation of clinical information to determine occurrence of adverse AEs. Another limitation of the study is that validation was constrained by a small sample size (50 notes total, double-evaluated), which was limited by the availability of annotators and the time-intensive nature of annotation. Low IRR for AEs nature may reflect differences in the evaluators’ training backgrounds. Class imbalance (e.g., fewer positive cases for some non-prespecified events) was another limitation, reflecting the lower likelihood of occurrence for certain AEs. Additionally, the model’s context-window limitations required truncation or chunking of longer notes, limiting our ability to assess performance on lengthy, longitudinal charts spanning multiple encounters and potentially obscuring cross-document dependencies. This limitation may be mitigated by newer models with substantially larger context windows than the gpt-4-0314 model used in this study, enabling more complete evaluation of longer clinical narratives and cross-note relationships. Non-prespecified AEs and clinical interventions were not embedded by any methods to reduce computational steps and minimize the time required for reviewers. Clustering these would enhance interpretation and highlights the need for further research on how to integrate this step. The study employed zero-shot prompting without additional training, context, or guidelines for the model, which may have impacted overall performance but provides a foundation for future methodological refinements, performance improvements and an important step in oncology for establishing the potential of LLMs to identify and extract CAR-T-related AEs from clinical notes. Moreover, we envision validating this method in other healthcare systems, which would aid in better interpretation of the results and broader and more accurate application.

## Conclusion

We applied and evaluated an LLM to analyze clinical notes and extract AEs, including CRS and ICANS presence, grades, dates, and related interventions, from unstructured progress notes. Performance was high for CRS and ICANS presence and grade, with strong to near-perfect inter-rater reliability, moderate-to-high performance for CRS and ICANS dates and interventions, and more variable performance for broader AE nature, date and intervention categories.

These results support the integration of LLM-derived extraction of CRS and ICANS grading into EHR workflows, safety monitoring platforms, epidemiology studies and real-world evidence pipelines, where working with unstructured data is time-consuming and timely CRS and ICANS grading and/or detection is critical.

Documentation variability and class imbalance for broader AEs remain challenges, highlighting the need for further research into LLM interpretation of clinical results Additionally, cautious use of LLMs in a zero-prompt manner for broad AE extraction is warranted due to the variable performance observed in this study. Nevertheless, this study provides a foundation for further research, including the introduction of LLMs into real-world evidence studies and safety monitoring in clinical trials and oncology practice, while ensuring that performance metrics are carefully monitored and interpreted.

## Acknowledgments

The authors thank the Butte Lab, including Ana Maria Deluca, Boris Oskotsky, Dana Ludwig, Harry Sun, Jaysón Davidson, Michelle Wang, Noah Baker, Sanchita Bhattacharya, Sirui Ding, Travis Zack, and Reuben Sarwal. We also extend our gratitude to the Sirota Lab led by Marina Sirota, including Laura Almonte, Umair Khan, and Brenda Ametepe. Additionally, we thank the Research Data Assets & Information Commons Team, the Research Infrastructure Team, Gerald (Gerry) Fontenay, and the staff of the UCSF Bakar Computational Health Sciences Institute. We are grateful for the support provided by UCSF-Stanford CERSI, including Kathy Giacomini and Lawrence Lin, as well as Nathan Dang, Holly Ly, and Jaime Kenyon. Finally, we acknowledge Gilead and Shahed Iqbal for their contributions and collaboration in JG fellowship.

## Author contributions

JG, BM, and AJB conceived the study. JG, BM designed the methodology. JG and BM curated the data. MSu, CYKW, RV, TTO, MSi, and AJB provided data resources and artificial intelligence expertise and insights. JG, AS, CYKW, and TTO provided clinical expertise and insights. JG and BM wrote the code. JG performed formal analyses. JG and AS evaluated LLM performance by reviewing the notes. JG prepared the tables and figures. MSi and AJB provided resources, supervision, and project administration. JG drafted the original manuscript, and all authors contributed to review and editing and approved the final version.

## Institutional Review Board Statement

Data used in this study was drawn from the UCSF Health system, where deidentification of the data has been completed to enable clinical research projects, under guidance from UCSF institutional review boards (IRBs), privacy, and compliance.

## Informed Consent Statement

Identifiable patient information was not used in this study.

## Data Availability Statement

The data supporting this study are restricted due to the sensitivity of deidentified patient data. Access is not available upon request.

