## Supplementary Materials for "Extracting adverse event nature, severity, timelines and resulting interventions from clinical notes of patients receiving CAR-T therapy using large language models"

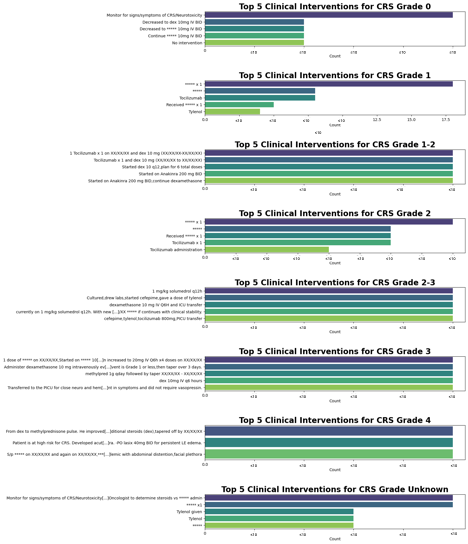

Figure S1 Frequencies of the most frequent resulting clinical intervention by CRS Grades

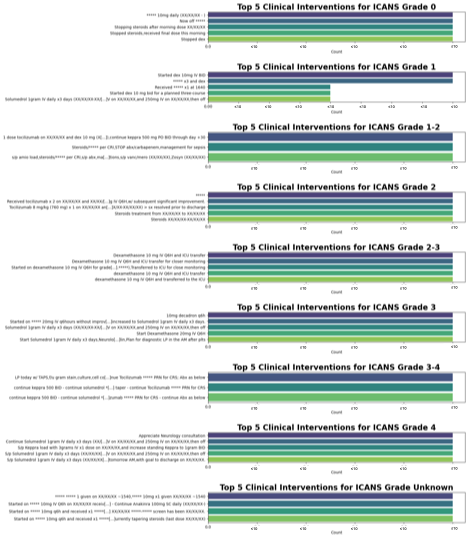

Figure S2 Frequencies of the most frequent resulting clinical intervention by ICANS Grades

Table S1 Prompts used for extracting: (i) prespecified adverse events, dates of occurrence, and clinical interventions; (ii) non-prespecified adverse events, their dates of occurrence, and clinical interventions; (iii) CRS occurrences, their grades, dates, and clinical interventions; and (iv) ICANS occurrences, their grades, dates, and clinical interventions.

prompt1 = """Task: Extract adverse events due to CAR-T administration, their dates (format mm-dd-yyyy), and the clinical interventions performed from the clinical note provided. Format the output in CSV format with the headers “Adverse event”, “Date of Adverse Event”, and “Clinical intervention”. Ensure that the “Adverse event” is one of the following values: “Cytokine Release Syndrome (CRS)”, “Immune Effector Cell-Associated Neurotoxicity Syndrome (ICANS)”, “Hypogammaglobulinemia”, “Neutropenic Fever”, “Macrophage Activation Syndrome (MAS)”, “Hemophagocytic Lymphohistiocytosis (HLH)”, “Tumor Lysis Syndrome”, “Movement disorder”, “Cognitive impairment”, “Personality changes”, “B-cell aplasia”, “Thrombocytopenia”, “Anemia”, “Neutropenia”, “Leukopenia”, “Hypogammaglobulinemia”, “CD4 lymphopenia”, “Cranial Nerve Palsy”, “Bacterial infection”, “Viral Infection”, “Zoster Infection”, “Fungal infection”, “Aspergillus infection”, “Pneumocystis Jirovecii Pneumonia infection”, “Secondary myeloid malignancies”, and “Graft-versus-host disease (GVHD)”. If no adverse event is identified, fill “Adverse event” with “No” and “NA” for “Date of Adverse Event” and “Clinical intervention”. If an adverse event is identified but no date or clinical intervention is provided, note “NA” for “Date of Adverse Event” or “Clinical intervention”. Information mentioned in the transfusion threshold is a general guideline for clinicians and might not reflect the occurrence of an adverse event or a clinical intervention."""

prompt2 = """Task: Extract adverse events due to CAR-T administration, their dates (format mm-dd-yyyy), and clinical interventions from the clinical note provided. Format the output in CSV format with the headers “Adverse event”, “Date of Adverse Event”, and “Clinical intervention”. Exclude adverse events with the following categories: “Cytokine Release Syndrome (CRS)”, “Immune Effector Cell-Associated Neurotoxicity Syndrome (ICANS)”, “Hypogammaglobulinemia”, “Neutropenic Fever”, “Macrophage Activation Syndrome (MAS)”, “Hemophagocytic Lymphohistiocytosis (HLH)”, “Tumor Lysis Syndrome”, “Movement disorder”, “Cognitive impairment”, “Personality changes”, “B-cell aplasia”, “Thrombocytopenia”, “Anemia”, “Neutropenia”, “Leukopenia”, “Hypogammaglobulinemia”, “CD4 lymphopenia”, “Cranial Nerve Palsy”, “Bacterial infection”, “Viral Infection”, “Zoster Infection”, “Fungal infection”, “Aspergillus infection”, “Pneumocystis Jirovecii Pneumonia infection”, “Secondary myeloid malignancies”, and “Graft-versus-host disease (GVHD)”. If no adverse event is identified, fill “Adverse Event” with “No” and “NA” for “Date of Adverse Event” and “Clinical intervention”. If an adverse event is identified but no date or clinical intervention is provided, note “NA” for “Date of Adverse Event” or “Clinical intervention”. Information mentioned in the transfusion threshold is a general guideline for clinicians and might not reflect the occurrence of an adverse event or a clinical intervention."""

prompt3 = """Task: Extract the occurrence of Cytokine Release Syndrome (CRS), the grade of CRS, and clinical interventions from the provided clinical note. Format the output in CSV format with the headers “CRS”, “CRS grade”, “Date of occurrence of CRS”, and “Clinical intervention”. If CRS is identified, fill “CRS” with “Yes”, “CRS grade” with the corresponding grade (format Grade X, Grade could be between 0 to 4), “Date of occurrence of CRS” with the date (format mm-dd-yyyy) of CRS, and “Clinical intervention” with the clinical intervention performed to treat the CRS. If there is no CRS, fill “CRS” with “No” and “NA” for the “CRS grade”, “Date of occurrence of CRS”, and “Clinical intervention”. If CRS is identified but no CRS grade, no date, or no clinical intervention is identified, note “Yes” for “CRS” and “NA” for the “CRS grade”, “Date of occurrence of CRS”, or “Clinical intervention”. Information mentioned in the transfusion threshold is a general guideline for clinicians and might not reflect the occurrence of CRS or a clinical intervention."""

prompt4 = """Task: Extract the occurrence of Immune Effector Cell-Associated Neurotoxicity Syndrome (ICANS), the grade of ICANS, and clinical interventions from the provided clinical note. Format the output in CSV format with the headers “ICANS”, “ICANS grade”, “Date of occurrence of ICANS”, and “Clinical intervention”. If ICANS is identified, fill “ICANS” with “Yes”, “ICANS grade” with the corresponding grade (format Grade X, Grade could be between 0 to 4), “Date of occurrence of ICANS” with the date (format mm-dd-yyyy) of ICANS, and “Clinical intervention” with the clinical intervention performed to treat the ICANS. If there is no ICANS, fill “ICANS” with “No” and “NA” for the “ICANS grade”, “Date of occurrence of ICANS”, and “Clinical intervention”. If ICANS is identified but no ICANS grade, no date, or no clinical intervention is identified, note “Yes” for “ICANS” and “NA” for the “ICANS grade”, “Date of occurrence of ICANS”, and “Clinical intervention”. Information mentioned in the transfusion threshold is a general guideline for clinicians and might not reflect the occurrence of ICANS or a clinical intervention."""

Table S2 Performances of the Large Language Models for each prompt by reviewer Jordan Guillot (JG) and Arvind Suresh (AS)

|  | JG |  |  |  | AS |  |  |  | JG |  |  |  | AS |  |  |  | Cohen’s kappa |
| --- | --- | --- | --- | --- | --- | --- | --- | --- | --- | --- | --- | --- | --- | --- | --- | --- | --- |
|  | True Positive | True Negative | False Positive | False Negative | True Positive | True Negative | False Positive | False Negative | Accuracy | Precision | Recall | F1 Score | Accuracy | Precision | Recall | F1 Score |  |
| Prompt Prespecified AE |  |  |  |  |  |  |  |  |  |  |  |  |  |  |  |  |  |
| Nature | 114 | 8 | 46 | 10 | 85 | 10 | 75 | 3 | 0.69 | 0.71 | 0.91 | 0.80 | 0.55 | 0.53 | 0.96 | 0.69 | 0.401 |
| Date | 86 | 62 | 45 | 10 | 90 | 70 | 41 | 3 | 0.73 | 0.66 | 0.90 | 0.76 | 0.78 | 0.69 | 0.97 | 0.80 | 0.545 |
| Clinical intervention | 97 | 29 | 66 | 1 | 114 | 32 | 50 | 6 | 0.65 | 0.60 | 0.99 | 0.74 | 0.72 | 0.70 | 0.95 | 0.80 | 0.448 |
| Prompt Non Prespecified AE |  |  |  |  |  |  |  |  |  |  |  |  |  |  |  |  |  |
| Nature | 21 | 32 | 10 | 11 | 18 | 41 | 13 | 2 | 0.72 | 0.68 | 0.66 | 0.67 | 0.80 | 0.58 | 0.90 | 0.71 | 0.606 |
| Date | 12 | 42 | 10 | 11 | 13 | 45 | 9 | 2 | 0.72 | 0.55 | 0.52 | 0.53 | 0.84 | 0.59 | 0.87 | 0.70 | 0.634 |
| Clinical intervention | 16 | 46 | 11 | 1 | 18 | 45 | 9 | 2 | 0.84 | 0.59 | 0.94 | 0.72 | 0.85 | 0.67 | 0.90 | 0.77 | 0.681 |
| Prompt CRS |  |  |  |  |  |  |  |  |  |  |  |  |  |  |  |  |  |
| Presence | 26 | 19 | 5 | 0 | 28 | 19 | 3 | 0 | 0.90 | 0.83 | 1.0 | 0.91 | 0.94 | 0.90 | 1.0 | 0.95 | 0.857 |
| Grade | 36 | 19 | 1 | 2 | 36 | 19 | 0 | 1 | 0.95 | 0.97 | 0.95 | 0.96 | 0.98 | 1.0 | 0.97 | 0.99 | 0.961 |
| Date | 46 | 19 | 10 | 2 | 47 | 19 | 8 | 2 | 0.84 | 0.82 | 0.96 | 0.88 | 0.89 | 0.85 | 0.96 | 0.90 | 0.777 |
| Clinical intervention | 38 | 19 | 9 | 2 | 44 | 19 | 2 | 2 | 0.84 | 0.80 | 0.95 | 0.87 | 0.94 | 0.96 | 0.96 | 0.96 | 0.771 |
| Prompt ICANS |  |  |  |  |  |  |  |  |  |  |  |  |  |  |  |  |  |
| Presence | 7 | 39 | 3 | 1 | 6 | 38 | 4 | 2 | 0.92 | 0.70 | 0.88 | 0.78 | 0.88 | 0.60 | 0.75 | 0.67 | 0.792 |
| Grade | 10 | 39 | 2 | 1 | 8 | 38 | 4 | 2 | 0.94 | 0.83 | 0.91 | 0.87 | 0.88 | 0.67 | 0.80 | 0.73 | 0.862 |
| Date | 15 | 39 | 23 | 1 | 16 | 38 | 22 | 2 | 0.69 | 0.39 | 0.94 | 0.56 | 0.69 | 0.42 | 0.89 | 0.57 | 0.919 |
| Clinical intervention | 10 | 39 | 6 | 1 | 13 | 38 | 3 | 2 | 0.88 | 0.63 | 0.91 | 0.74 | 0.91 | 0.81 | 0.87 | 0.84 | 0.777 |

Table S3 Top 50 resulting clinical intervention for prespecified adverse events

| **Clinical intervention** | **Frequency (patients)** | **%** |
| --- | --- | --- |
| No transfusions needed today | 48 | 16.40% |
| Tocilizumab administration | 46 | 15.70% |
| Transfuse for Hb<7,plt<10k | 25 | 8.50% |
| Cefepime administration | 18 | 6.10% |
| Assess and record max CRS twice/day. If CRS develops,increase lab frequency to daily. | 17 | 5.80% |
| Continue levaquin and fluconazole until no longer neutropenic | 17 | 5.80% |
| Assess and record max ICANS score daily and prn. RN neuro checks q shift. | 17 | 5.80% |
| Transfusion Threshold: Hgb < 7,Plt < 10 | 16 | 5.50% |
| Administered ***** | 14 | 4.80% |
| Administered cefepime | 14 | 4.80% |
| Administered ***** x 1 | 13 | 4.40% |
| Transfuse to keep hgb>7 | 13 | 4.40% |
| Transfuse to keep plt >10 | 13 | 4.40% |
| Received ***** x 1 | 12 | 4.10% |
| Transfusion: Hgb >8,Platelet >10 | 12 | 4.10% |
| Administration of tocilizumab | 11 | 3.80% |
| Dexamethasone administration | 11 | 3.80% |
| No clinical intervention mentioned | 11 | 3.80% |
| No transfusions or cytokines needed today | 11 | 3.80% |
| No transfusions or cytokines needed today. | ≤10 | ≤3.4% |
| s/p IV Zosyn XX/XX/XX - XX/XX/XX in setting of neutropenic fevers | ≤10 | ≤3.4% |
| IV Zosyn XX/XX/XX - XX/XX/XX | ≤10 | ≤3.4% |
| No transfusions needed today. | ≤10 | ≤3.4% |
| Monitoring for signs/symptoms of CRS/Neurotoxicity | ≤10 | ≤3.4% |
| No specific intervention mentioned | ≤10 | ≤3.4% |
| Monitor for signs/symptoms of CRS/Neurotoxicity | ≤10 | ≤3.4% |
| Administered ***** x1 | ≤10 | ≤3.4% |
| Administration of ***** x1 | ≤10 | ≤3.4% |
| Platelet transfusion | ≤10 | ≤3.4% |
| Monitoring for fever,hypotension | ≤10 | ≤3.4% |
| Ceftriaxone administration | ≤10 | ≤3.4% |
| Transfuse PRBCs for Hgb<7 (or sx) or with bleeding using CMV negative,irradiated,and leukoreduced products. Premed with tylenol and atarax 10mg po | ≤10 | ≤3.4% |
| Patient at high risk for developing CRS or neurotoxicity | ≤10 | ≤3.4% |
| Started on cefepime | ≤10 | ≤3.4% |
| Started cefepime | ≤10 | ≤3.4% |
| Administered ***** and dexamethasone | ≤10 | ≤3.4% |
| Tocilizumab | ≤10 | ≤3.4% |
| Transfuse to keep hgb>7,plt >10 | ≤10 | ≤3.4% |
| Given ***** x1 | ≤10 | ≤3.4% |
| No transfusions today | ≤10 | ≤3.4% |
| Clinical monitoring | ≤10 | ≤3.4% |
| tocilizumab | ≤10 | ≤3.4% |
| No transfusions needed | ≤10 | ≤3.4% |
| Started on ***** 10mg IV Q6h on XX/XX/XX received a total of 3 doses then increased to 20mg IV Q6h doses on XX/XX/XX-> switched to Solumedrol 1000mg due to severe bradycardia XX/XX/XX dex (XX/XX/XX,XX/XX/XX,XX/XX/XX) | ≤10 | ≤3.4% |
| Transfuse with CMV negative,LR,IR products. For platelet transfusion needs acetaminophen,diphenhydramine,and famotidine for premedication; has history of hives with transfusions. No premeds with blood. Avoid hydrocortisone. | ≤10 | ≤3.4% |
| Heme goals HgB >7,plts >10K | ≤10 | ≤3.4% |
| Tylenol given | ≤10 | ≤3.4% |
| Cefepime 2g IV Q8h | ≤10 | ≤3.4% |
| Monitor for signs/symptoms of CRS/Neurotoxicity - Fever/Hypotension/Tachycardia/Hypoxia - Mental status changes - RN to cont BID neuro checks per study protocol - Oncologist to determine steroids vs ***** admin - Page fellow *****-***** with CRS/Neurotox symptoms | ≤10 | ≤3.4% |
| Cefepime 2g IV q8h | ≤10 | ≤3.4% |

Table S4 Category of Non-Prespecified Adverse Events as describe in the results

ae_categories = {

"Fever & Inflammatory Responses": [

"Fever", "Febrile", "Intermittent fevers", "Persistent fevers",

"Rigors", "Mild chills", "Fevers and tachycardia",

"Cytokine Release Syndrome (CRS)", "Grade 4 CRS",

"Recurrent fever", "Transient fever", "Febrile overnight",

"Low grade temp 99.3", "Fever and neutropenia",

"Febrile and tachycardic", "Transient rigors",

"Neutropenic fever", "Elevated inflammatory markers",

"Febrile to Tmax 39.3C overnight",

"Mild fever",

"Shivering",

"Rising CRP",

"Intermittent fevers + hypotension to 72/46"

],

"Gastrointestinal Issues": [

"Nausea", "Nausea and vomiting", "Diarrhea", "Watery diarrhea",

"Loose stool", "Toxic enterocolitis", "Esophagitis",

"Dyspepsia/bloating/gas", "Severe constipation",

"C difficile infection", "Toxic Gastroenteritis", "Proctitis",

"Abdominal pain", "Diffuse abdominal pain", "GI bleed",

"Lower GI bleed", "Acute diarrhea",

"Hemorrhagic UGIB", "Constipation", "Severe nausea",

"Persistent diarrhea", "Enterocolitis due to C-Diff infection",

"Toxic enteritis", "Bloody diarrhea ? hematuria",

"Diarrhea, resolved", "Intermittent diarrhea",

"Abdominal tenderness", "Toxic enterocolitis secondary to chemotherapy",

"Dysphagia", "Nausea/mild epigastric discomfort",

"Solid-food dysphagia", "Chronic diarrhea",

"Severe diarrhea", "Constipation-resolved", "Enterocolitis due to chemotherapy",

"Abdominal pain and rectal pain", "Toxic gastroenteritis",

"Diffuse abdominal tenderness", "Stercoral-colitis",

"Recurrent small bowel obstruction", "Acute pancreatitis",

"Dysgeusia", "Toxic Enteritis",

"Chronic Diarrhea",

"Abdominal Pain",

"Recurrent GI Bleed",

"Diarrhea",

"Diarrhea,resolved",

"Diarrhea/loose stools",

"Odynophagia",

"Ischemic colitis",

"Acalculous cholecystitis",

"Pelvic abscess",

"Prolapse",

"Contained perforation adjacent to the anastomosis",

"Small abscesses",

"Hemorrhagic cystitis",

"Rectal pain",

"Severe Constipation",

"Abd pain and rectal pain",

"LLQ abdominal pain",

"Mild constipation",

"New abdominal tenderness",

"Soft stools",

"Esophageal HSV",

"Altered bowel habits",

"Severe dysphagia and severe protein/calorie malnutrition",

"Hemorrhoids",

"Constipation,resolved",

"Acinetobacter UTI c/b Hematuria from R nephrostomy tube",

"Nausea & vomiting",

"Nausea/vomiting",

"Nausea and Vomiting",

"Nausea and emesis",

"Nausea/Vomitting",

"Emesis with scant blood tinge",

"Acute on chronic extremity swelling due to volume overload",

"Pneumoperitoneum",

"Possibly cholecystitis",

"Elevated LFT",

"Gingival bleeding",

"Vaginal bleeding",

"Non-gap metabolic acidosis, improving",

"Shock Liver",

"Poor PO intake",

"Malnutrition",

"Volume overload",

"*****: Hemorrhagic UGIB",

"Metabolic acidosis",

"Orbital cellulitis",

"Oral thrush",

"Hypofibrinogenemia",

"Acid reflux",

"Hyperbilirubinemia",

"Transaminitis",

"Cholestatic/hepatocellular liver injury",

"Nausea/Vomiting",

"Elevated Transaminases",

"Worsening LFTs",

"Low Fibrinogen",

"Abnormal brain CT",

"Aspiration event",

"Dyspepsia",

"Anorexia, Dysgeusia (taste alteration), Nausea/Vomiting",

'Elevated LFTs', 'abdominal Pain', 'diarrhea', 'Intermittent nausea', 'Severe abdominal pain',

'GERD', 'IBS/GERD', 'History of pancreatitis', 'Disorder of nutrition',

'Hemorrhagic Chemosis', 'Anorexia,Dysgeusia (taste alteration),Nausea/Vomitting',

'Glucose trending up as steroids restarted', 'UGIB - resolved', 'UGIB',

"Hiccups",

"Emesis",

"N/V",

'transaminitis' ,

"LFT elevation",

],

"Neurological Issues and Dizziness": [

"Acute encephalopathy", "Neurotoxicity", "Altered mental status",

"Confusion", "Headache", "Persistent chronic headache", "Seizure activity",

"Status epilepticus", "Migraine", "Mild neuropathy", "Neuropathy",

"Grade 4 Neurotoxicity", "Cognitive impairment", "Personality changes",

"Movement disorder", "Visual disturbances", "Retinal Hemorrhages",

"Migraine and blackout vision", "Optic nerve edema",

"Memory loss", "Unsteady gait", "Mild neurotoxicity",

"Acute encephalopathy, possible seizure", "Acute visual disturbance",

"Brain fog", "New AMS overnight", "Acute Encephalopathy 2/2 Grade 2 Neurotoxicity",

"Seizure", "Seizure like activity", "Essential Tremor vs myoclonus",

"Hypnopompic hallucinations", "Slow speech and takes time to find words",

"Difficulty choosing the right words", "Transient vertigo",

"Facial neuro deficits", "Acute stroke", "New ischemic stroke",

"Migraine and black out vision", "Acute metabolic encephalopathy",

"Acute stroke, small", "Worsening mental status",

"Severe headache", "Hand tremor", "Dizziness,Hypotension",

"Brain AVF", "Acute encephalopathy,hand tremor",

"Acute encephalopathy,possible seizure", "Acute Retinal Hemorrhages",

"Persistent headache", "New hallucinations", "Hallucination",

"Seizures", "Symptomatic orthostasis", "Neuropathic pain",

"Visual floaters in bilateral eyes", "Binocular Diplopia",

"Headache,progressive", "Numbness/Twitching in hands and feet",

"Blurred vision", "Acute distress","Confusion,gait changes,and mild bilateral hand tremors",

"Impaired Vision",

"Decrease in hearing",

"Encephalopathy: Metabolic evidenced by delirium",

"Encephalopathy and BLE weakness",

"Encephalopathy due to *****-T neurotoxicity,delirium",

"Toxic Encephalopathy",

"At risk for CRS/Neurotoxicity",

"Grade 2 Encephalopathy",

"Neurotoxicity req steroids",

"Developing hydrocephalus",

"Obstructive hydrocephalus",

"Chronic trichiasis",

"Episode of blacked out vision with headache",

"AMS",

"Persistent nausea",

"Seizure like activity/encephalopathy",

"Grade ***** *****",

"Chorioretinopathy,bilateral",

"Vision loss",

"Permanent optic nerve damage",

"Right sided facial droop",

"R sided facial droop",

"Minor tremor in hands",

"Essential tremor",

"Stable hand tremors",

"Increased tremor",

"Hand tremors",

"Headaches",

"Mild headache",

"Ocular headache",

"Somnolence",

"Late morning drowsiness",

"Sleeping trouble",

"Syncopal episode",

"Syncopal event",

"Dizziness",

"Dizziness with ambulation",

"Orthostatic dizziness",

"Dizziness,risk of *****",

"Dizziness with standing",

"Symptomatic orthostatic hypotension",

"Orthostatic hypotension/dizziness",

"Deconditioned",

"Deconditioned from prolonged/multiple hospital stays",

"Hypoxemia - intermittent",

"High risk for falls",

"Blacked out vision and headache",

"Left chin numbness",

"L chin numbness",

"Pathologic L humeral fracture",

"Splenic infarct",

"Intermittent shin pain",

"Dizziness", "Lightheadedness", "Loss of consciousness",

"Dizziness with ambulation", "Orthostatic dizziness", "Dizziness & orthostasis",

"Lightheadedness after standing", "Dizziness and feeling high after low dose marinol",

"Symptomatic orthostatics",

"Orthostatic dizziness",

"Symptomatic orthostatic hypotension",

"Syncope",

"Delirium",

"At risk for CRS/Neurotoxicity",

"Tinnitus",

"Possible seizure",

"Acute stroke, small",

"Encephalopathy",

"Peripheral neuropathy",

"Severe agitation",

"Encephalitis",

"CRS/neurotoxicity",

"Vertigo",

"Metabolic Encephalopathy",

"Treating encephalopathy",

"Numbness in feet",

"Left shoulder and bicep pain",

"Syncopal episode with head trauma",

"Pain: R neck and shoulder pain from neck mass",

"Cognition",

"Generalized body aches",

"Full body aches",

"Restless leg syndrome",

"Pain of left thigh",

"Peripheral neuropathy due to chemotherapy",

"Chronic neuropathic pain",

"Left Upper Extremity Pain",

"Throat tightness and swelling under tongue",

"Intermittent hematuria",

'Acute stroke,small', 'Left foot drop', 'Subdural hematoma', 'Chronic cancer-related pain',

'Vocal cord atrophy/paralysis of LEFT true vocal cord', 'Post-herpetic neuralgia', 'Cauda equina syndrome',

'Seizure XX/XX/XX lymphoma\xa0in ***** XX/XX/XX, 'Mild occipital headache',

'Encephalopathy: Toxic Encephalopathy', 'Non-infectious SIRS due to *****-T cell therapy with End ***** Damage present (toxic encephalopathy)',

'Patient passed out', 'Bilateral visual loss from optic nerve atrophy due to leukemic involvement',

'Back pain', 'Restless Legs', 'Pain at port site', 'Chronic cancer pain', 'R>L shoulder pain',

"Vision loss of right eye",

"Blurry vision",

"Right frontal extra-axial collection",

"Mild PN",

"***** changes",

"Hx SDH",

"SDH"

],

"Cardiovascular Issues": [

"Hypertension", "Hypotension", "Orthostatic hypotension",

"Orthostatic tachycardia", "Bradycardia", "Sinus bradycardia",

"Tachycardia", "Prolonged QTC", "Afib with RVR",

"Paroxysmal Atrial Fibrillation", "VTach overnight",

"Saddle Pulmonary Embolus", "Orthostatic HoTN", "Sinus tachycardia",

"Atrial Tachycardia", "Transient Afib w/ RVR", "Paroxysmal SVT",

"A flutter", "Prolonged QTc", "Mild persistent neuropathy",

"Transient ***** potentially c/w grade 2 neurotoxicity",

"Tachy-brady", "Sinus Tachycardia", "New onset Afib",

"Paroxysmal Atrial Fibrillation c/b RVR",

"Low Blood Pressure",

"Hypotension",

"Orthostatic hypotention",

"Orthostatic Hypotension",

"Hypotensive to 82/49",

"hypotensive to 72/46",

"Sinus Bradycardia",

"A fib with RVR",

"Afib w/ RVR",

"Atrial tachycardia",

"VTach overnight and 6 beats this AM",

"Multiple PVCs",

"4 seconds atrial tach",

"QTc prolonged",

"Risk of QTc prolongation",

"Prolonged Qtc",

"Prolonged PT",

"High D-dimer",

"Elevated d-dimer",

"Tachycardia up to 130 with ambulation",

"Tachycardia / anxiety",

"Heartburn",

"Chest pain",

"Chest pain during ***** infusion",

"Chest Pain/SOB",

"Chest Pain/SOB/Orthostasis",

"Chest Pain/SOB/*****",

"Acute Saddle PE",

"PE",

"Incidental PE",

"Hx of Pulmonary embolism/SMV thrombus",

"Hx of Pulmonary embolism/SMV thrombus s/p anticoagulation",

"Saddle PE",

"Multiple pulmonary emboli",

"Pericardial effusion",

"Pericardial effusion ?malignant",

"Possibly left ovarian vein thrombus",

"Nonocclusive right jugular venous thrombosis",

"Non-occlusive right axillary DVT",

"Non-occlusive DVT in R axillary vein",

"Non-occlusive DVT of RUE",

"DVT",

"DVT of RLE",

"LLE DVT",

"RUE DVT",

"R IJ Port associated thrombus",

"R malignant pleural effusion",

"Bilateral pleural effusions",

"Bilateral pleural effusions R>L ?malignant",

"CKD stage III",

"Hx of incidental PE",

"hx of incidental PE",

"Hx of ARF",

"HTN",

"OSA",

"Septic shock",

"Mild pulmonary edema",

"Risk for vision loss",

"Hemoptysis",

"Hypoxemia",

"Hypercarbia",

"R pleural effusion",

"Hx of HTN",

"Hx of paroxysmal SVT",

"Hx of AT",

"Bradycardia during ***** infusion",

"Non ischemic cardiomyopathy",

"HTN established, controlled",

"Hypotensive",

"Orthostasis",

"Aflutter",

"Paroxysmal Atrial Fibrillation c/b RVR now resolved",

"Chronic Sinus bradycardia",

"Asymptomatic Baseline Hypotension",

"Prior CDI",

'hx of HTN', 'Rule out PE', 'At risk for CRS/ Neurotoxicity', 'Hx of possible pericardial involvement by lymphoma',

'VTE', 'Hypertension,controlled', 'HTN established,controlled', 'HTN,Established,uncontrolled',

'Chronic RIJ Occlusive thrombus', 'H/o Splenic infarct', 'Bilateral pleural effusions R>L\xa0?malignant',

'Pericardial effusion\xa0?malignant', 'Low BP', 'VTE PPx: Contraindicated: CNS mets',

'Asymptomatic Bradycardia', 'intermittent fevers + hypotension to 72/46', 'At risk for CRS/Neurotoxicity after *****-T infusion',

'HCM', 'Acute on chronic subdural bleed', 'Capillary leak', 'R IJ/R Subclavian DVT related to ***** catheter',

'LLE DVT s/p IVC filter', 'Deep Vein Thrombosis', 'Methemoglobinemia',

'Chronic RIJ Occlusive thrombus', 'H/o Splenic infarct',

"***** w/ RVR overnight",

"***** w/ RVR",

"Hx of HFpEF",

"Hx DVT",

"Hx of incidental PE",

],

"Respiratory Issues": [

"Dyspnea", "Hypoxia", "Acute hypoxic respiratory failure",

"Respiratory distress", "Pulmonary edema", "Asthma",

"Obstructive Hydrocephalus", "Pneumocystis Jirovecii Pneumonia infection",

"Pulmonary aspergillosis", "Hypoxic respiratory failure",

"Acute dyspnea", "Acute hypoxia", "Respiratory failure",

"Respiratory failure: Acute with hypoxemia", "Multifocal GGOs, C/f PNA",

"Pulmonary infection", "Oxygen requirement",

"Increased dyspnea on exertion and increased fatigue",

"Respiratory distress in setting of fluid overload",

"Cough,Rhinovirus +", "Dyspnea on exertion",

"Shortness of breath",

"Hypoxia - intermittent",

"Acute on chronic hypoxic respiratory failure",

"***** hypoxic respiratory failure",

"Acute on chronic hypoxic respiratory failure",

"Acute Saddle PE","Cough",

"OSA treated with BiPAP",

"Obstructive sleep apnea",

"Bilateral pleural effusions R>L ?malignant",

"Pericardial effusion ?malignant",

"Multifocal GGOs, C/f PNA",

"c/f PNA",

'Increased dyspnea (now at rest) and ***** ***** ***** ***** *****% while sitting',

'Obstructive sleep apnea on CPAP', 'Severe OSA', 'Multifocal GGOs,C/f PNA', 'Sinus infection',

'Sinus headaches', 'Bilateral conjunctivitis', 'Bilateral conjunctivitis/Hemorrhagic Chemosis',

'Mild edema of epiglottis and arytenoids',

"Sore throat"

],

"Renal & Electrolyte Issues": [

"Acute kidney injury (AKI)", "AKI on CKD",

"Acute kidney injury on CKD III", "Microscopic hematuria",

"Hyperkalemia", "Hypokalemia", "Hyponatremia", "Hypercalcemia",

"Low Urea Nitrogen", "Elevated Creatinine", "Renal function impairment",

"Hematuria", "Persistent lactic acidosis", "Non-gap metabolic acidosis",

"Non-gap metabolic acidosis, improving", "AKI due to postobstructive uropathy",

"Hypomagnesemia", "Low Potassium", "High Chloride", "Low CO2",

"Low Calcium", "Low Albumin", "Low Phosphorus",

"Acute kidney injury on CKD III", "Acute hyponatremia",

"Renal Failure", "AKI,due to postobstructive uropathy",

"Increased Creatinine", "Acute Kidney Injury",

"Acute kidney injury (AKI)", "AKI (Established,improved)",

"Renal abscess", "AKI due to interval worsening B hydronephrosis due to malignancy",

"Recurrent AKI","AKI",

"Acute Kidney Injury",

"Acute Kidney Injury (AKI)",

"Acute kidney injury",

"Hx of ARF",

"Non-gap metabolic acidosis, improving",

"Hyperkalemia",

"Mild Hyponatremia",

"Polyuria",

"Urinary urgency",

"Urinary hesitancy",

"AKI (baseline Cr ~1)",

"Cr elevated to 1.50",

"Hypoalbuminemia",

"R ureteral obstruction",

"AKI, R ureteral obstruction",

'ARF', 'Cr elevated-1L NS IVF bolus given', 'HYPERkalemia', 'Urinary retention', 'CKD',

'Low Uric Acid', 'Low Creatine kinase', 'Low ALT', 'High Direct Bilirubin', 'High Ferritin',

'Low Abs Neutrophils', 'Low Abs Lymphocytes', 'Low Abs Monocytes', 'Low Fibrinogen,Functional',

'High Fibrin D-Dimers', 'Hypernatremia', 'Urinary frequency', 'CKD XX/XX/XX renal lymphoma involvement',

'Chronic Kidney Disease: CKD Stage IIIa', 'AKI,R ureteral obstruction', 'Cystitis symptoms',

'Non-gap metabolic acidosis,improving',

"CKD stage",

"Hyperlipidemia",

"Hyperlipidemia, controlled",

'Hyperlipidemia,controlled',

"Hx of hyperuricemia",

"Incontinence",

'Urology - increased nocturia'

],

"Infections": [

"Bacterial infection", "Viral infection", "Fungal infection",

"Zoster Infection", "Staph epi bacteremia", "Pseudomonas bacteremia",

"Strep Mitis bacteremia", "Acinetobacter UTI", "C diff colitis",

"Vaginitis", "Urine Cx: >100,000 Staph Epi", "Odontogenic infection",

"COVID infection", "COVID Viral infection", "COVID-19 infection",

"COVID-19 infection, recovered", "RSV", "HHV-6 infection",

"Ecoli bacteremia", "ESBL UTI", "Stomatitis, presumed fungal",

"Vaginitis with vaginal white papules", "Sepsis", "Septal resection site with fungal elements",

"Skin infection at PICC site", "Fungemia", "Septic embolization",

"Achromobacter bacteremia", "Acute COVID infection",

"Non-neutropenic fever", "Non-neutropenic fever,suspected pulmonary source (resolved)",

"COVID + infection", "Strep mitis bacteremia",

"Rhino positive", "Septic shock due to GNR bacteria",

"Rhinovirus infection",

"COVID Infection",

"COVID Viral infection",

"COVID-19 infection,recovered",

"Staph epi central line infection/colonization",

"C. diff",

"C. Diff colitis",

"CDiff colitis",

"Recurrence of C. Diff colitis",

"Odontogenic infection",

"MDR pseudomonas and E faecium complicated UTI",

"C.diff",

"Klebsiella CLABSI/bacteroides fragilis bacteremia",

"Orbital cellulitis",

"UTI",

"GPC bacteremia",

"Cervical adenopathy (Left cervical neck mass)",

"Stomatitis, presumed fungal",

"External otitis media",

"Cytomegalovirus (CMV) viremia",

"Hx Osteomyelitis s/p resection and abx spacers",

"Osteomyelitis",

"Hx of COVID Viral infection",

"L axillary wound",

'Orbital cellulitis R>L', 'hx of Oral thrush', 'c/f SSTI / superinfection of chest wall mass',

'Reported hx of PSC,Established,controlled', 'H/o COVID Infection', 'Treated for sepsis',

'Hematuria,resolved', 'Stomatitis,presumed fungal', 'Sinus infection',

"Prostatic abscess",

"Hepatitis B core antibody positive",

"Complex liver cyst with hemorrhage"

],

"Hematologic & Oncologic Issues": [

"Pancytopenia", "Cytopenias", "Neutropenia",

"Hemophagocytic Lymphohistiocytosis (HLH)",

"Macrophage Activation Syndrome (MAS)", "Tumor Lysis Syndrome",

"Refractory B-Cell Lymphoma", "Malignancy associated fatigue",

"Secondary myeloid malignancies", "Anemia associated with malignancy",

"Neoplasm/malignancy associated pain", "Pain due to leukemia",

"Malignancy-associated pain", "Cancer Related Pain", "Relapsed diffuse large B-cell lymphoma",

"Thrombocytopenia secondary to malignancy", "Bone disease",

"Advanced Care Planning", "Relapsed DLBCL",

"B-cell ALL,s/p XX/XX/XX MUD BMT XX/XX/XX",

"Refractory *****-Cell Lymphoma", "Pancytopenia due to chemotherapy",

"Functional SBO due to abdominal lymphomatosis",

"Recurrent malignant ascites", "Primary refractory high-grade B-cell lymphoma (double hit) with CNS involvement",

"Large B-cell Lymphoma,Stage IV,Refractory",

"Disease associated pancytopenia", "Profound thrombocytopenia",

"Pancytopenia XX/XX/XX disease", "Multiple Myeloma",

"Chronic Neuropathy", "Neoplasm/malignancy associated fatigue",

"Pain,Malignancy-associated",

"pancytopenia",

"Cytopenia",

"Anemia due to malignancy",

"Cancer-related pain",

"Malignancy associated fatigue on admission",

"Relapsed DLBCL,non-GCB",

"Refractory DLBCL",

"B-cell ALL",

"Coagulopathy",

"At risk of tumor lysis syndrome",

"Possible pericardial involvement by lymphoma",

"Adrenal insufficiency (CMS code)",

"Acute lymphoblastic leukemia",

"Possible MDS",

"Proliferative Diabetic Retinopathy",

"Platelet refractory",

"Pancytopenia due to lymphodepleting chemotherapy",

"Profound leukocytosis",

"Thyroid nodule",

"Cancer Pain",

"Severe protein-calorie malnutrition",'Low WBC Count', 'Low Hemoglobin', 'Low Hematocrit', 'Low RBC Count', 'Low Platelet Count',

'Low ***** Count,Flow Cytometry', 'High PT', 'High INR', 'High aPTT', 'Athlete\'s foot',

'***** ALL (acute lymphoblastic leukemia)', 'ALL (acute lymphoblastic leukemia)',

'Non-infectious SIRS due to *****-T cell therapy', 'Throat itching', 'Failure to thrive,malnutrition',

'Bilateral foveal subretinal fluid/serous detachment,consistent with very mild central serous retinopathy',

'Abnormal mandibular marrow signal adjacent to left trigeminal nerve in the mandibular foramen',

'Lytic bone disease',

"LT groin ulcer",

"Hx R humerus fracture"

],

"Skin & Soft Tissue Issues": [

"Rash", "Papular rash", "Skin Rash", "Pruritis", "Conjunctivitis",

"Diffuse Body Erythema", "RUE swelling and pruritis",

"Upper back erythema", "Intertrigo", "Diffuse Erythema",

"Erythematous papular rash to face", "Rash on back",

"Pruritic back rash", "Rash on arms, back", "Skin changes over hands",

"Maculopapular rashes", "Full body rash", "Bleeding from back",

"Radiation dermatitis", "Non-infectious SIRS due to supraglottic arytenoid and epiglottic edema",

"New rash", "Skin lesions", "Rash present",

"Maculopapular rashes on the chest and forehead",

"Rash associated with CAR-T infusion", "Rash on legs and hands",

"Diffuse skin changes", "Erythematous follicular papules","Rash on arms,back",

"Skin rash",

"Diffuse skin changes",

"Pruritus",

"Dry skin over hands/feet",

"Dry skin/dry mouth",

"Dry eye",

"Abrasion on the right chest from previous dressing",

"Dried blood noted",

"Facial/eye swelling",

"Submandibular swelling",

"Submandibular gland swelling",

"Ingrown toenail",

"Rash, resolving",

"Gluteal muscle soreness",

"Bilateral eye swelling",

"Perirectal abrasions",

"L axillary wound",

"Right first toe injury with scar formation",'***** lower extremity pain', 'L groin pain', 'Achy', 'Hypothermia', 'Rash,resolving',

'Swelling (right hand is swollen,not erythematous,non tender. R arm with edema that is also not tender.)',

'Cervical adenopathy ( Left cervical neck mass)', 'Superficial venous thrombosis', 'Left upper chest pain',

'Small amount of bleeding at port site',

"***** swelling",

"LT groin ulcer"

],

"Musculoskeletal Issues": [

"Bone pain", "Back pain", 'back pain', "Chronic Back pain", "Shoulder pain",

"L Ankle pain", "LLE weakness/pain", "Sacral pain due to herniated disc",

"Pain in legs and hips", "New onset back pain", "Diffuse Cerebral Microhemorrhages",

"Pathologic right femoral neck fracture", "Impending Left humeral fracture",

"Painful sensory neuropathy and tremors", "Severe sinus Bradycardia",

"Left sided TMJ pain", "R hip replacement", "Shoulder pain",

"Severe R temporal pain", "Pain management", "Left shoulder pain",

"Neck and R hip pain", "L Groin pain", "Back/left hip pain",

"Bilateral LE neuropathy", "RUE pain", "R lower cheek melanoma",

"Pain (primarily back and leg)", "Right shoulder pain",

"Acute back pain", "Bone pain: low back,hips,legs","Musculoskeletal Issues",

"Chronic pain",

"Chronic back pain",

"Low back pain",

"Back pain due to radiculopathy",

"Muscle cramping",

"Muscle weakness",

"Generalized pain to chest wall",

"Weakness in the legs",

"Right foot 5th metatarsal fracture",

"Hand cramping",

"Pain of left thigh",

"Joint pain",

"Left shoulder strain",

"Supraspinatus tear and tendinosis",

"Multiple vertebral osseous lesions",

"Frequent falls",

"Physical Deconditioning",

"Back pain",

"Bilateral foot neuropathy",

"Fall",

'L lower extremity pain', 'Crampy muscle spasms', 'Persistent left shoulder and bicep pain',

'Bilateral feet neuropathy', 'Right hand index finger fracture',

'Impaired mobility',

"Gout flair",

"Gout flair R hallux",

"Back pain",

"Hx of R humerus fracture"

],

"Psychiatric & Behavioral Issues": [

"Anxiety", "Depression", "Agitation", "Panic attack", "Insomnia",

"Increased somnolence", "Hallucinations", "Personality changes",

"Depression/anxiety", "Adjustment disorder w/ anxiety",

"Severe Depression", "Depressed mood and affect",

"Significant fatigue", "Anxiety about health",

"Major depressive disorder,single episode,moderate",

"H/o Passive Suicidal Ideation-resolved", "Anxiety/insomnia",

"Insomnia; Adjustment disorder w/ anxiety","Psych",

"Anxiety",

"Agitation",

"Depression",

"Adjustment disorder",

"Anxiety/Insomnia",

"Hx of Depression",

"Severe agitation",

"Restless leg syndrome",

'Pain', 'Drenching night sweats', 'Chills', 'Vasovagal episode',

"*****'s Disease"

],

"Endocrine Issues": [

"Adrenal insufficiency", "Diabetes Mellitus (T2DM)", "Hyperglycemia",

"Hypoglycemia", "Steroid-induced hyperglycemia", "Steroid-induced hypertension",

"Partial adrenal insufficiency", "Steroid induced hyperglycemia",

"Fasting hyperglycemia", "Secondary adrenal insufficiency",

"Steroid-induced hypertension", "Not well controlled DMII","Diabetes mellitus",

"Refractory hypoglycemia",

"Adrenal Insufficiency",

"Steroid-induced side effects",

"Hypothyroidism",

"Hx of Hypoglycemia, DM2",

"Adrenal *****",

"Hypoalbuminemia",

'Hypothyroid', 'H/o Hypoglycemia,DM2', 'T2DM', 'DM: Established,controlled'

],

"Immune System Issues": [

"Immunocompromised", "B-cell aplasia", "Graft-versus-host disease (GVHD)",

"CD4 lymphopenia", "Hypogammaglobulinemia", "Immunodeficiency",

"Functional immunosuppression", "Anaphylactic shock",

"Immunocompromised state", "Immune-related fatigue","Immunocompromised State",

"Neutropenic precautions",

"High C-Reactive Protein",

"Cancer related pain",

"Perineural spread of tumor",

'At risk for CRS/ Neurotoxicity', 'At risk of CRS/Neurotoxicity', '*****-T therapy requiring close monitoring for CRS/neurotoxicity',

"Allergies"

],

"Medication Side Effects": [

"Cefepime toxicity", "Steroid-induced side effects",

"Chemotherapy-related pancytopenia", "Toxic enteritis secondary to chemotherapy",

"Anaphylaxis", "Allergic reaction to Cefepime",

"Toxic gastroenteritis of chemotherapy", "Chemotherapy induced pancytopenia",

"Chemotherapy induced nausea and vomiting", "Toxic Enteritis","Methotrexate toxicity",

"Opioid induced constipation",

"Treating encephalopathy with increased dex",

"Decreased mealtime insulin",

'Glucose trending up as steroids restarted', 'Methemoglobinemia'

],

"Fatigue and weakness": [

"Fatigue", "Weakness", "Generalized weakness, fatigue and nausea",

"Exhaustion", "Chronic fatigue", "Fatigue/Dizziness",

"Deconditioning", "Fatigue/weakness", "Profound fatigue",

"Fatigue and Lethargy", "Extreme fatigue with cytoxan",

"General fatigue", "Muscle aches,profound fatigue,insomnia,bruising and anxiety",

"Acute on Chronic Fatigue,Generalized Weakness", "Low energy",

"Low energy level",

"Generalized weakness,fatigue and nausea",

"Fatigue",

"Generalized body aches",

"Full body aches",

"Ill-appearing",

'Fatigue and weakness', 'Weight loss', 'Weight gain',

"Mild pain near the kidney regions",

"Mild *****"

],

"Appetite loss and poor oral intake": [

"Appetite loss", "Poor oral intake",

"Decreased appetite", "Anorexia", "Early satiety", "Loss of appetite",

"Low appetite", "Poor appetite", "Decreased oral intake",

"Low appetite,weight loss",

"Decreased appetite/weight loss",

"Poor PO intake",

"Malnutrition",

"Lower appetite",

"Failure to thrive, malnutrition",

"Nutritional assessment",

'Insufficient calorie intake', 'Failure to thrive,malnutrition'

],

"Edema": [

"Edema", "Bilateral LE edema", "LLE Edema", "Trace edema to BLE's with L >R",

"Bilateral pleural effusions R>L", "Orbital edema", "Scrotal Edema",

"Anasarca", "Anasarca RUE", "Anasarca RUE with pitting edema",

"Edema in Right lower leg", "R hand swelling", "R arm swelling",

"Bilateral pleural effusions R>L",

"Scrotal,LE edema",

"Anasarca",

"Anasarca RUE with pitting edema but other extremities with no pitting edema but do appear to gradually swell",

"Bilateral edema",

"Mild fluid overload",

"Swelling (right hand is swollen, not erythematous, non tender. R arm with edema that is also not tender.)",

"Leg edema",

"Acute on chronic extremity swelling", 'RUE swelling', 'L arm swelling and discomfort distal to PICC', 'Bilateral pleural effusions R>L\xa0?malignant'

],

"Other AEs": [

"Elderly", "Hx weight loss", "Social", "Advanced Care Planning",

"Complex sx management", "Requires assistance with activities of rehabilitation",

"Developmental delay", "Distributive shock",

"Disease progression", "Nutritional Assessment",

"Fluid/Electrolytes", "Bone health", "Obesity",

"Fluid overload",

"Fluid status: Volume overload",

"Volume overload requiring IV diuresis",

"Protein calorie malnutrition due to SBO",

"No",

"Glaucoma",

"HLD",

"S/p cataract surgery L eye",

"Hair thinning",

"Sensitivity at tongue",

"Bowel and bladder incontinence",

"Episode of fecal incontinence",

'??cecal mass', 'Parotitis', 'BPH established,controlled', 'BPH', 'Hearing Changes', 'Tooth pain',

'Pain at port site',

"Nan",

"nan",

np.nan

]

}

Table S5 Top 50 Clinical Intervention for Non-Prespecified Adverse Events

| **Clinical intervention** | **Frequency (patients)** | **Percent** |
| --- | --- | --- |
| CTM | ≤ 10 | ≤ 3.4% |
| Correct with bicarbonate supplementation in ***** | ≤ 10 | ≤ 3.4% |
| Received third-line R-***** C1D1 XX/XX/XX | ≤ 10 | ≤ 3.4% |
| Continuous tele | ≤ 10 | ≤ 3.4% |
| Zofran RTC,compazine,Ativan PRN | ≤ 10 | ≤ 3.4% |
| Lidocaine patch to R shoulder,Oxycodone 2.5mg q6h PRN,PT/OT | ≤ 10 | ≤ 3.4% |
| Currently on IV abx | ≤ 10 | ≤ 3.4% |
| Zofran,compazine,Ativan PRN | ≤ 10 | ≤ 3.4% |
| Senna 17.2mg PO BID,Imodium prn,Miralax prn | ≤ 10 | ≤ 3.4% |
| Started on H1/H2 blockade,unasyn,vancomycin | ≤ 10 | ≤ 3.4% |
| Hospitalization | ≤ 10 | ≤ 3.4% |
| Cardiology consult,Monitor closely on telemetry,Stop offending medications,including zofran and dex,Steroids now switched from ***** to Solumedrol,although solumedrol can also cause bradycardia,Changed Fluc to caspo on XX/XX/XX | ≤ 10 | ≤ 3.4% |
| Empiric treatment for esophageal HSV,Fluconazole,Nystatin | ≤ 10 | ≤ 3.4% |
| dex 10mg q 6 hrs ( XX/XX/XX-XX/XX/XX)--> 10mg q 8hr (XX/XX/XX) --> 10mg q 12 (XX/XX/XX- ) | ≤ 10 | ≤ 3.4% |
| Now off ***** | ≤ 10 | ≤ 3.4% |
| Senna,Imodium,Miralax | ≤ 10 | ≤ 3.4% |
| CTM,Avoiding zofran,Use steroids cautiously if needed | ≤ 10 | ≤ 3.4% |
| Increased levetiracetam to 1500 mg BID | ≤ 10 | ≤ 3.4% |
| Midodrine 5 mg PO TID,1L NS bolus x1,mIVF @ 100ml/hr | ≤ 10 | ≤ 3.4% |
| Dexamethasone administration | ≤ 10 | ≤ 3.4% |
| Sent to ED for infectious workup | ≤ 10 | ≤ 3.4% |
| Increased levetiracetam 1500 mg BID | ≤ 10 | ≤ 3.4% |
| Apply a thin layer of Mupirocin to purulent areas,other areas please cover with vaseline,then apply ABD pad,then secure with paper tape BID | ≤ 10 | ≤ 3.4% |
| Urgent rad onc consult scheduled for XX/XX/XX at ***** | ≤ 10 | ≤ 3.4% |
| continue Gabapentin 400mg qAM and qafternoon,800mg qHS | ≤ 10 | ≤ 3.4% |
| Started dex 10mg IV BID | ≤ 10 | ≤ 3.4% |
| Completed dex 10mg IV BID until XX/XX/XX PM and switched to 5mg IV BID starting XX/XX/XX | ≤ 10 | ≤ 3.4% |
| Continue dex 10mg IV BID | ≤ 10 | ≤ 3.4% |
| Continue dex 10mg IV TID | ≤ 10 | ≤ 3.4% |
| Removal of PICC,Hold further AC given thrombocytopenia | ≤ 10 | ≤ 3.4% |
| CMP daily,consider renal consult if worsens | ≤ 10 | ≤ 3.4% |
| Start Solumedrol 1gram IV daily x3 days | ≤ 10 | ≤ 3.4% |
| Continue physiologic replacement: -cortef 10mg qAM - 5mg q Afternoon - 5mg qPM -stress dose: hydrocortisone 50 mg/m2 = 100 mg,followed by 12.5 mg/m2 (26mg) q6 hours x 24 hours | ≤ 10 | ≤ 3.4% |
| Encourage PO intake | ≤ 10 | ≤ 3.4% |
| Hold fluconazole,using micafungin for ppx | ≤ 10 | ≤ 3.4% |
| Stopped metop on XX/XX/XX | ≤ 10 | ≤ 3.4% |
| Increase Midodrine to 5mg PO TID | ≤ 10 | ≤ 3.4% |
| Added flomax | ≤ 10 | ≤ 3.4% |
| Hold fluconazole,using micafungin for ppx,d/c-ed tylenol on XX/XX/XX | ≤ 10 | ≤ 3.4% |
| Stop metformin | ≤ 10 | ≤ 3.4% |
| Lung expansion pathway with RT | ≤ 10 | ≤ 3.4% |
| Developed acute hypoxia on XX/XX/XX,requiring 6L NC. Hypotension on XX/XX/XX responsive to IV fluids. CT chest XX/XX/XX consistent with pulmonary edema. | ≤ 10 | ≤ 3.4% |
| Start Midodrine 2.5mg PO TID | ≤ 10 | ≤ 3.4% |
| Started on dex,received 10mg IV @ 1839 | ≤ 10 | ≤ 3.4% |
| Cont vimpat 100mg BID for seizure ppx,continue salt tabs 3g TID,further adjustments pending sodium trend; fluid restriction lifted as it was limiting his solute intake | ≤ 10 | ≤ 3.4% |
| Received 1 dose of ***** | ≤ 10 | ≤ 3.4% |
| Foley placement | ≤ 10 | ≤ 3.4% |
| Continues on ***** 10mg IV TID | ≤ 10 | ≤ 3.4% |
| Cardiology consult,Atropine at bedside,Monitor closely on telemetry,Stop offending medications,including zofran and dex,Steroids now switched from ***** to Solumedrol,Changed Fluc to caspo,daily ECG | ≤ 10 | ≤ 3.4% |
| Added flomax XX/XX/XX | ≤ 10 | ≤ 3.4% |

Table S6 Top 30 Clinical Intervention for Grade 0 CRS

| **Clinical intervention** | **Frequency (patients)** |
| --- | --- |
| Monitor for signs/symptoms of CRS/Neurotoxicity | ≤ 10 |
| Decreased to dex 10mg IV BID | ≤ 10 |
| Decreased to ***** 10mg IV BID | ≤ 10 |
| Continue ***** 10mg IV BID | ≤ 10 |
| No intervention | ≤ 10 |
| Notify heme fellow or attending for fevers,hemodynamic instability,hypoxia,or neuro changes. Heme attending to decide if tocilizumab is appropriate if c/f CRS. Infectious work up and treatment PRN for fevers (including any neutropenic fever). Monitor ICE score per protocol. | ≤ 10 |
| No new interventions | ≤ 10 |
| No signs of CRS or neurotoxicity since. | ≤ 10 |
| No steroids | ≤ 10 |
| None so far | ≤ 10 |
| Notify heme fellow or attending for fevers,hemodynamic instability,hypoxia,or neuro changes. Heme attending to decide if tocilizumab is appropriate if c/f CRS. Infectious work up and treatment PRN for fevers (including any neutropenic fever). Monitor ICE score per protocol CRS. | ≤ 10 |
| ***** 10mg IV BID | ≤ 10 |
| Now off ***** | ≤ 10 |
| Now off dex | ≤ 10 |
| Off dex | ≤ 10 |
| RN to cont BID neuro checks per study protocol | ≤ 10 |
| Received dex 10 mg IV | ≤ 10 |
| Received dex 10mg | ≤ 10 |
| Received dex 10mg IV | ≤ 10 |
| Received tocilizumab #2,dex 10mg x1 | ≤ 10 |
| Notify heme fellow or attending for fevers,hemodynamic instability,hypoxia,or neuro changes; Heme attending to decide if tocilizumab is appropriate if c/f CRS; Infectious work up and treatment PRN for fevers (including any neutropenic fever); Monitor ICE score per protocol | ≤ 10 |
| No decadron | ≤ 10 |
| Received tylenol x3 | ≤ 10 |
| Increased dexamethasone to 10mg q6H | ≤ 10 |
| Heme attending ***** ***** ***** tocilizumab is appropriate if c/f CRS | ≤ 10 |
| ICE XX/XX/XX | ≤ 10 |
| ICE pending admission | ≤ 10 |
| Increased ***** to 10mg q6H | ≤ 10 |
| Increased ***** to 10mg q6H (XX/XX/XX-XX/XX/XX) | ≤ 10 |
| Increased dex 10mg IV to q8h | ≤ 10 |

Table S7 Top 30 Clinical intervention for Grade 1 CRS

| **Clinical intervention** | **Frequency (patients)** |
| --- | --- |
| ***** x 1 | 18 |
| ***** | ≤ 10 |
| Tocilizumab | ≤ 10 |
| Received ***** x 1 | ≤ 10 |
| Tylenol | ≤ 10 |
| ***** x1 | ≤ 10 |
| s/p ***** x 1 | ≤ 10 |
| Oncologist to determine steroids vs ***** admin | ≤ 10 |
| tocilizumab | ≤ 10 |
| 2nd dose of ***** | ≤ 10 |
| Tocilizumab x 1 | ≤ 10 |
| ***** x 1 XX/XX/XX at 0309 then second dose XX/XX/XX at 1734 | ≤ 10 |
| One dose of ***** | ≤ 10 |
| Resolved with ***** | ≤ 10 |
| ***** given x 1 | ≤ 10 |
| ***** given x1 | ≤ 10 |
| Given ***** | ≤ 10 |
| tocilizumab x 1 | ≤ 10 |
| Received ***** | ≤ 10 |
| Tocilizumab administration | ≤ 10 |
| s/p ***** | ≤ 10 |
| Monitor for signs/symptoms of CRS/Neurotoxicity,RN to cont BID neuro checks per study protocol,Oncologist to determine steroids vs ***** admin | ≤ 10 |
| Monitor for signs/symptoms of CRS/Neurotoxicity | ≤ 10 |
| Received ***** x 1 on XX/XX/XX | ≤ 10 |
| Administered ***** and dex | ≤ 10 |
| ICE XX/XX/XX | ≤ 10 |
| Tylenol given | ≤ 10 |
| Tocilizumab #1 | ≤ 10 |
| Tocilizumab dose #1 | ≤ 10 |
| Tocilizumab administration on XX/XX/XX,Supportive care with Tylenol q6h PRN,Continuous pulse ox | ≤ 10 |

Table S8 Top 30 clinical intervention for Grade 1-2 CRS

| **Clinical intervention** | **Frequency (patients)** |
| --- | --- |
| 1 Tocilizumab x 1 on XX/XX/XX and dex 10 mg (XX/XX/XX-XX/XX/XX) | ≤ 10 |
| Tocilizumab x 1 and dex 10 mg (XX/XX/XX to XX/XX/XX) | ≤ 10 |
| Started dex 10 q12,plan for 6 total doses | ≤ 10 |
| Started on Anakinra 200 mg BID | ≤ 10 |
| Started on Anakinra 200 mg BID,continue dexamethasone | ≤ 10 |
| Started on Anakinra 200 mg BID,increased dex to 10mg Q6H | ≤ 10 |
| Tocilizumab administration,Dex administration | ≤ 10 |
| Tocilizumab administration,Dexamethasone administration | ≤ 10 |
| Tylenol given for persistent fevers & tachycardia,developing pain | ≤ 10 |
| 1 dose of tocilizumab,dex 10 mg Q12h,zosyn,keppra for prophy | ≤ 10 |
| Tylenol given. ***** given for persistent fevers & tachycardia,developing pain | ≤ 10 |
| Tylenol given. ***** given for persistent fevers & tachycardia,developing pain. EKG,echo,CXR done for c/o lower sternal pain | ≤ 10 |
| Tylenol,***** | ≤ 10 |
| s/p ***** | ≤ 10 |
| s/p *****,amiodarone gtt,metoprolol IV pushes | ≤ 10 |
| tocilizumab (ACTEMRA) IVPB (adult),steroids/***** per CRI,STOP abx/carbapenem,management for sepsis | ≤ 10 |
| Started dex 10 q12 | ≤ 10 |
| Spiked a fever,hypertension. Cultures,VBG,viral w/u ordered. Tylenol given. ***** given for persistent fevers & tachycardia,developing pain. EKG,echo,CXR done for c/o lower sternal pain. C/o headaches and lower back pain (respond to tylenol and dilaudid). ALC 90. | ≤ 10 |
| Spiked a fever,hypertension. Cultures,VBG,viral w/u ordered. Tylenol given | ≤ 10 |
| S/p ***** x 1 (XX/XX/XX,XX/XX/XX),dex 10mg x1 (XX/XX/XX),then dex 10mg q6H (XX/XX/XX-XX/XX/XX),10 mg q12 (XX/XX/XX-XX/XX/XX),then stopped | ≤ 10 |
| Received 2 doses of ***** and started on Decadron 10mg PO BID | ≤ 10 |
| Increased to 10mg Q6H | ≤ 10 |
| Increased dex to 10mg Q6H | ≤ 10 |
| Dexamethasone treatment | ≤ 10 |
| Decadron 10mg PO BID,***** x2 | ≤ 10 |
| Continued Anakinra 200 mg BID,increased dex to 10mg Q6H | ≤ 10 |
| Continued Anakinra 200 mg BID,continued dex | ≤ 10 |
| CTM,Page CRI for any new concerns for CRS | ≤ 10 |
| Amio load,steroids/***** per CRI,management for sepsis,IVF,Afib control: Amio gtt to PO (XX/XX/XX- ) with goal to load 4-6g (will receive 2.5g by IV in first 24hr) Metop tartrate 25 q6h [ ] back up- esmolol gtt > dilt gtt,Pressors PRN: 1st line levophed,***** for CRS--> decadron started and to continue per CRI,APAP for fevers,***** repletions | ≤ 10 |

TableS9 Top 30 Clinical Intervention for Grade 2 CRS

| **Clinical intervention** | **Frequency (patients)** |
| --- | --- |
| ***** x 1 | ≤ 10 |
| ***** | ≤ 10 |
| Received ***** x 1 | ≤ 10 |
| Tocilizumab x 1 | ≤ 10 |
| Tocilizumab administration | ≤ 10 |
| Tocilizumab and Dexamethasone administration | ≤ 10 |
| Dexamethasone 10mg IV BID (XX/XX/XX-) | ≤ 10 |
| Treated with ***** | ≤ 10 |
| ***** x2 | ≤ 10 |
| ***** x 1 on XX/XX/XX | ≤ 10 |
| ***** x1 | ≤ 10 |
| Tocilizumab | ≤ 10 |
| tocilizumab | ≤ 10 |
| Third dose of ***** administered,Restarted dexamethasone 10mg IV Q6H | ≤ 10 |
| Third dose of ***** administered on XX/XX/XX at 5:42 am | ≤ 10 |
| Oxygen and ***** x 2 | ≤ 10 |
| Sent infectious work up,started cefepime and received ***** x 1 | ≤ 10 |
| Second dose of Tocilizumab given | ≤ 10 |
| SBP <100,not responsive to fluids - given ***** x1 10am dose #1 - CRS G2 - ICE 10,no neurotox. 2100 - SBP <100,no further fevers,ordered ***** dose #2 - discussed with RN - discussed with night float - message heme fellow on call Monitor VS closely through the night. Consider Dose #3 around 0500 (8hrs from last dose) based upon VS. | ≤ 10 |
| S/p ***** x1 on XX/XX/XX @0730,s/p ***** x1 on XX/XX/XX@***** | ≤ 10 |
| S/p ***** x1 on XX/XX/XX @0730 | ≤ 10 |
| S/p ***** x 1 dose and dex 10 mg IV Q6H on XX/XX/XX. ***** deescalated to Q8H on XX/XX/XX. ***** deescalated to Q12H on XX/XX/XX. Received addition dose on XX/XX/XX. Last day of dex on XX/XX/XX. | ≤ 10 |
| S/p ***** on XX/XX/XX,dex 10mg x1 on XX/XX/XX,XX/XX/XX | ≤ 10 |
| Oxygen and ***** x 2 (XX/XX/XX & XX/XX/XX),Midodrine to 5mg PO TID (XX/XX/XX- ,last incr XX/XX/XX- ),Keppra 500mg BID through Day 30 (can stop after XX/XX/XX) | ≤ 10 |
| S/p ***** on XX/XX/XX,S/p ***** on XX/XX/XX,S/p dex x4 doses for CRS/neurotox | ≤ 10 |
| S/p ***** on XX/XX/XX,S/p ***** on XX/XX/XX,Monitor for symptoms of CRS/neurotoxicity,Keppra 500mg BID ppx | ≤ 10 |
| S/p ***** on XX/XX/XX and again on XX/XX/XX - Weaned off oxygen XX/XX/XX. | ≤ 10 |
| S/p ***** on XX/XX/XX and XX/XX/XX,dex 10mg x1 on XX/XX/XX,XX/XX/XX | ≤ 10 |
| Restarted dexamethasone 10mg IV Q6H | ≤ 10 |
| Restarted Dexamethasone 10mg IV BID on XX/XX/XX and now with somewhat improving mental status,and concern for underlying CRS/neurotoxicity. Decrease Dexamethasone to 10mg IV q12 (XX/XX/XX--> ) (previously on XX/XX/XX 10 BID,XX/XX/XX 10q6,XX/XX/XX 10 TID) | ≤ 10 |

Table S10 All Clinical Intervention for Grade 2-3 CRS

| **Clinical intervention** | **Frequency (patients)** |
| --- | --- |
| 1 mg/kg solumedrol q12h | ≤ 10 |
| Cultured,drew labs,started cefepime,gave a dose of tylenol | ≤ 10 |
| dexamethasone 10 mg IV Q6H and ICU transfer | ≤ 10 |
| currently on 1 mg/kg solumedrol q12h. With new onset facial tingling,MRI brain w/wo and ***** demonstrated multiple osseous lesions at calvarium and skull base with increased enhancement. Started solumedrol wean to 0.5mg/kg q12h on XX/XX/XX ***** if continues with clinical stability. | ≤ 10 |
| cefepime,tylenol,tocilizumab 800mg,PICU transfer | ≤ 10 |
| cefepime,tylenol,NS 1L bolus,***** 800mg | ≤ 10 |
| Tylenol,***** x1,NS bolus,albumin,methylpred started. Third spacing. Seen by PICU/RRT team but supportive care measures had good responses and vitals stabilized. Lasix 10mg given once vitals stabilize to diurese some fluid | ≤ 10 |
| Tylenol,***** x1,NS bolus,albumin,methylpred started. Lasix 10mg given once vitals stabilize to diurese some fluid | ≤ 10 |
| Tylenol,***** x1,NS bolus,albumin,methylpred started | ≤ 10 |
| Tylenol,***** x1,NS bolus,albumin,methylpred | ≤ 10 |
| On 1 mg/kg solumedrol q12h. Started solumedrol wean to 0.5mg/kg q12h on XX/XX/XX | ≤ 10 |
| On 1 mg/kg solumedrol q12h,started solumedrol wean to 0.5mg/kg q12h on XX/XX/XX | ≤ 10 |
| On 1 mg/kg solumedrol q12h | ≤ 10 |
| NS 1L bolus given followed by a dose of ***** 800mg and PICU made aware | ≤ 10 |
| Febrile,tachycardia,hypotensive,hypoxic. Tylenol,***** x1,NS bolus,albumin,methylpred started. Third spacing. Seen by PICU/RRT team but supportive care measures had good responses and vitals stabilized. Lasix 10mg given once vitals stabilize to diurese some fluid. Pain other than intermittent headaches resolved,using dilaudid prn. ALC 160. Discussed with Dr. *****. ***** dose repeated at 2230 for fevers and hypotension. | ≤ 10 |
| Febrile,tachycardia,hypotensive,hypoxic. Tylenol,***** x1,NS bolus,albumin,methylpred started | ≤ 10 |
| Dexamethasone 10 mg IV Q6H and ICU transfer | ≤ 10 |
| Dexamethasone 10 mg IV Q6H | ≤ 10 |
| Currently on 1 mg/kg solumedrol q12h. With new onset facial tingling,MRI brain w/wo and ***** demonstrated multiple osseous lesions at calvarium and skull base with increased enhancement. Started solumedrol wean to 0.5mg/kg q12h on XX/XX/XX ***** if continues with clinical stability. | ≤ 10 |
| Cultured,drew labs,started cefepime,gave a dose of tylenol. Transferred to PICU following 2nd bolus when BP dropped to 70s/40s | ≤ 10 |
| Cultured,drew labs,started cefepime,gave a dose of tylenol. Discussed with Dr. *****,IT attending. Pt was persistently and increasingly febrile and tachycardic with decreased BP XX/XX/XX and 1 episode of hypoxemia 95% at approx. 14:30 -> NS 1L bolus given followed by a dose of ***** 800mg and PICU made aware. Transferred to PICU following 2nd bolus when BP dropped to 70s/40s as well as previously receiving prbc. No pressors required overnight,***** not needing redose. | ≤ 10 |
| Cultured,drew labs,started cefepime,gave a dose of tylenol. Discussed with Dr. *****,IT attending. Pt was persistently and increasingly febrile and tachycardic with decreased BP XX/XX/XX and 1 episode of hypoxemia 95% at approx. 14:30 -> NS 1L bolus given followed by a dose of ***** 800mg and PICU made aware. | ≤ 10 |
| Cultured,drew labs,started cefepime,gave a dose of tylenol,NS 1L bolus given followed by a dose of ***** 800mg and PICU made aware | ≤ 10 |
| methylprednisolone | ≤ 10 |

Table S11 All Clinical Intervention for Grade 3 CRS

| **Clinical intervention** | **Frequency (patients)** |
| --- | --- |
| 1 dose of ***** on XX/XX/XX,Started on ***** 10mg IV Q6h on XX/XX/XX received a total of 3 doses then increased to 20mg IV Q6h x4 doses on XX/XX/XX | ≤ 10 |
| Administer dexamethasone 10 mg intravenously every 6 hours. Continue dexamethasone use until the event is Grade 1 or less,then taper over 3 days. | ≤ 10 |
| methylpred 1g qday followed by taper XX/XX/XX - XX/XX/XX | ≤ 10 |
| dex 10mg IV q6 hours | ≤ 10 |
| Transferred to the PICU for close neuro and hemodynamic monitoring. S/p tocilizumab x1 and started methylprednisolone on XX/XX/XX with improvement in symptoms and did not require vasopressin. | ≤ 10 |
| Tocilizumab | ≤ 10 |
| Started on Decadron 20mg IV q6h,Stat NCHCT,Start EEG monitoring,Increase Keppra to 1,000mg IV BID,Blood and urine culture,Neurology consult | ≤ 10 |
| Started on Decadron 20mg IV q6h stat after discussion with attending. See other neuro eval/interventions below. Approximately 1 hour after first dose of steroids administered (~1100) pt with dramatic improvement,ICE score 10,remains disoriented but now forming sentences. Throughout the day patient with ongoing intermittent,aphasia,but overall improving encephalopathy. | ≤ 10 |
| Started on Decadron 20mg IV q6h | ≤ 10 |
| Started on ***** 10mg IV Q6h on XX/XX/XX received a total of 3 doses then increased to 20mg IV Q6h doses on XX/XX/XX. Solumedrol 1000mg x1 given XX/XX/XX. | ≤ 10 |
| Started on ***** 10mg IV Q6h on XX/XX/XX received a total of 3 doses then increased to 20mg IV Q6h doses on XX/XX/XX. Solumedrol 1000mg x1 given XX/XX/XX,XX/XX/XX. Ordered to receive again on XX/XX/XX. Continue Anakinra 100mg SC daily (XX/XX/XX-) | ≤ 10 |
| Started on ***** 10mg IV Q6h on XX/XX/XX received a total of 3 doses then increased to 20mg IV Q6h doses on XX/XX/XX | ≤ 10 |
| Start Dexamethasone 20mg IV Q6H | ≤ 10 |
| Received both ***** and Methylprednisolone | ≤ 10 |
| Received Tocilizumab x3 (XX/XX/XX,XX/XX/XX,XX/XX/XX) and started on Dexamethasone 10 mg IV BID for 3 days (XX/XX/XX-XX/XX/XX) | ≤ 10 |
| Dexamethasone 10mg IV q6h,Tocilizumab 8mg/kg IV | ≤ 10 |
| Continued dexamethasone 10mg IV q6h,tocilizumab administered on XX/XX/XX | ≤ 10 |
| Continue decadron 10mg q6h | ≤ 10 |
| Administer methylprednisolone 1 mg/kg intravenously twice daily or equivalent dexamethasone (e.g.,10 mg intravenously every 6 hours). Continue corticosteroids use until the event is Grade 1 or less,then taper over 3 days. | ≤ 10 |
| s/p ***** 10mg IV x 1 and x3 on XX/XX/XX | ≤ 10 |

Table S12 All Clinical Intervention for CRS Grade 4

| **Clinical intervention** | **Frequency (patients)** |
| --- | --- |
| From dex to methylprednisone pulse. He improved starting XX/XX/XX and nearly cleared by XX/XX/XX but recurred on XX/XX/XX and treated with additional steroids (dex),tapered off by XX/XX/XX | ≤ 10 |
| Patient is at high risk for CRS. Developed acute hypoxia on XX/XX/XX,requiring 6L NC. Hypotension on XX/XX/XX responsive to IV fluids. CT chest XX/XX/XX consistent with pulmonary edema. - S/p ***** on XX/XX/XX and again on XX/XX/XX - Weaned off oxygen XX/XX/XX. - please contact the CRI fellow (*****-*****) if patient is febrile,hypotensive,tachycardic,hypoxic or has changes in ***** or neurotoxicity signs - Ordered CT Chest w/o contrast on XX/XX/XX given persistent oxygen requirement - IV Lasix 40 mg x1 ordered on XX/XX/XX given patient appears hypervolemic with abdominal distention,facial plethora. -PO lasix 40mg BID for persistent LE edema. | ≤ 10 |
| S/p ***** on XX/XX/XX and again on XX/XX/XX,***** oxygen prn,CPO monitoring,Ordered CT Chest w/o contrast on XX/XX/XX given persistent oxygen requirement,IV Lasix 40 mg x1 ordered on XX/XX/XX given patient appears hypervolemic with abdominal distention,facial plethora | ≤ 10 |

Table S13 Top 30 Clinical Intervention for Grade 0 ICANS

| **Clinical intervention** | **Frequency (patients)** |
| --- | --- |
| ***** 10mg daily (XX/XX/XX - ) | ≤ 10 |
| Now off ***** | ≤ 10 |
| Stopping steroids after morning dose XX/XX/XX | ≤ 10 |
| Stopped steroids,received final dose this morning | ≤ 10 |
| Stopped dex | ≤ 10 |
| Started on dexamethasone 10mg IV q12h | ≤ 10 |
| Start midodrine 2.5mg TID | ≤ 10 |
| S/p Solumedrol 1gram IV daily x3 days (XX/XX/XX-XX/XX/XX) given highest c/f neurotoxicity,now on taper to 500mg IV on XX/XX/XX,and 250mg IV on XX/XX/XX,then off - Decadron 10mg q12h (XX/XX/XX - ) - S/p decadron 10mg q12h (XX/XX/XX - XX/XX/XX),s/p dex 10mg q6h (XX/XX/XX - XX/XX/XX) - Appreciate Neurology consultation - S/p Keppra load with 3grams IV x1 dose on XX/XX/XX,and increase standing Keppra to 1gram BID - OK to deescalate abx and defer LP - TTE,tele for stroke w/u negative - Delirium precautions - PT/OT/ST eval. PT eval'd XX/XX/XX w/ rec for home with PT | ≤ 10 |
| Remains off ***** | ≤ 10 |
| Remained off dex | ≤ 10 |
| Received ***** x1 at 1640 | ≤ 10 |
| RN neuro checks q shift | ≤ 10 |
| Now off dex | ≤ 10 |
| Notify Immunotherapy Attending | ≤ 10 |
| Taper ***** 10mg IV q12h | ≤ 10 |
| None so far | ≤ 10 |
| No steroids | ≤ 10 |
| No new interventions | ≤ 10 |
| Neurology consulted for new myoclonic activity EEG XX/XX/XX. Consider imaging if symptoms progress or EEG is abnormal. Continue neuro checks Continue Keppra for seizure ppx,start with chemo and continue until day +28 | ≤ 10 |
| Monitoring for signs/symptoms of CRS/Neurotoxicity,Levetiracetam 500mg BID for seizure prophylaxis | ≤ 10 |
| Monitoring for early neurologic signs,RN aware and monitoring closely | ≤ 10 |
| Monitor for signs/symptoms of CRS/Neurotoxicity,RN to cont BID neuro checks per study protocol,Cont Keppra,Oncologist to determine steroids vs ***** admin | ≤ 10 |
| Monitor for signs/symptoms of CRS/Neurotoxicity | ≤ 10 |
| Levetiracetam for seizure ppx,through Day 30 | ≤ 10 |
| Keppra started on first day of ***** chemotherapy and will continue through Day 21 post ***** infusion. | ≤ 10 |
| Keppra for seizure ppx,start Day 0 with ***** chemo and continue until day +28 | ≤ 10 |
| Supportive care and monitor neurologic status | ≤ 10 |
| Taper ***** 20mg IV q12h | ≤ 10 |
| Keppra for seizure ppx | ≤ 10 |
| dex 10mg x1 | ≤ 10 |

Table S14 Top 30 Clinical Intervention for Grade 1 ICANS

| **Clinical intervention** | **Frequency (patients)** |
| --- | --- |
| Started dex 10mg IV BID | ≤ 10 |
| ***** x3 and dex | ≤ 10 |
| Received ***** x1 at 1640 | ≤ 10 |
| Started dex 10 mg bid for a planned three-course | ≤ 10 |
| Solumedrol 1gram IV daily x3 days (XX/XX/XX-XX/XX/XX),now on taper to 500mg IV on XX/XX/XX,and 250mg IV on XX/XX/XX,then off | ≤ 10 |
| S/p Solumedrol 1gram IV daily x3 days (XX/XX/XX-XX/XX/XX) given highest c/f neurotoxicity,now on taper to 500mg IV on XX/XX/XX,and 250mg IV on XX/XX/XX,then off - Decadron 10mg q12h (XX/XX/XX - ) - S/p decadron 10mg q12h (XX/XX/XX - XX/XX/XX),s/p dex 10mg q6h (XX/XX/XX - XX/XX/XX) - Appreciate Neurology consultation - S/p Keppra load with 3grams IV x1 dose on XX/XX/XX,and increase standing Keppra to 1gram BID - OK to deescalate abx and defer LP - TTE,tele for stroke w/u negative - Delirium precautions - PT/OT/ST eval. PT eval'd XX/XX/XX w/ rec for home with PT | ≤ 10 |
| S/p Solumedrol 1gram IV daily x3 days (XX/XX/XX-XX/XX/XX) given highest c/f neurotoxicity,now on taper to 500mg IV on XX/XX/XX,and 250mg IV on XX/XX/XX,then off - Decadron 10mg BID (XX/XX/XX - ) - Appreciate Neurology consultation - S/p Keppra load with 3grams IV x1 dose on XX/XX/XX,and increase standing Keppra to 1gram BID - OK to deescalate abx and defer LP - TTE,tele for stroke w/u negative - Delirium precautions - PT/OT/ST eval,need ARU placement and will depend on neurotoxicity status | ≤ 10 |
| S/p Solumedrol 1gram IV daily x3 days (XX/XX/XX-XX/XX/XX) given highest c/f neurotoxicity,now on taper to 500mg IV on XX/XX/XX,and 250mg IV on XX/XX/XX,then off | ≤ 10 |
| S/p Keppra load with 3grams IV x1 dose on XX/XX/XX,and increase standing Keppra to 1gram BID | ≤ 10 |
| Resumed on decadron 10mg BID (XX/XX/XX - XX/XX/XX). | ≤ 10 |
| Received dex 10mg IV | ≤ 10 |
| Received dex 10mg | ≤ 10 |
| Received Tocilizumab x3 (XX/XX/XX,XX/XX/XX,XX/XX/XX) and started on Dexamethasone 10 mg IV BID for 3 days (XX/XX/XX-XX/XX/XX) | ≤ 10 |
| ***** 10mg IV x 1 given | ≤ 10 |
| Started dex 10mg IV BID,increased to 10mg x3 on XX/XX/XX,plan to taper to 10mg IV BID,then to 10mg IV daily,then to 4mg PO daily,then off | ≤ 10 |
| No new interventions | ≤ 10 |
| Methylpred 1mg/kg on XX/XX/XX | ≤ 10 |
| Managed as specified in overview | ≤ 10 |
| Keppra for seizure prophylaxis,started with ***** chemo and continued until day +21 (stopped XX/XX/XX) | ≤ 10 |
| Keppra for seizure prophylaxis | ≤ 10 |
| Increased to dex 10mg IV increased to q8h | ≤ 10 |
| Increased dex 10mg IV increased to q8h | ≤ 10 |
| Increased decadron 10mg q6h (XX/XX/XX - ) | ≤ 10 |
| Improving on ***** | ≤ 10 |
| None so far | ≤ 10 |
| Started dex 10mg IV q6h (for borderline G2 ICANs without concurrent CRS) | ≤ 10 |
| IV Dexamethasone 10mg q6h | ≤ 10 |
| Started dex,received 10mg IV | ≤ 10 |
| tapering dex,now 20mg IV q8h | ≤ 10 |
| s/p solumedrol 1g IV daily x 3 doses (XX/XX/XX-XX/XX/XX) then 250mg IV x 1 on XX/XX/XX | ≤ 10 |

Table S15 All Clinical Intervention for Grade 1-2 ICANS

| **Clinical intervention** | **Frequency (patients)** |
| --- | --- |
| 1 dose tocilizumab on XX/XX/XX and dex 10 mg (XX/XX/XX-XX/XX/XX),continue keppra 500 mg PO BID through day +30 | ≤ 10 |
| Steroids/***** per CRI,STOP abx/carbapenem,management for sepsis | ≤ 10 |
| s/p amio load,steroids/***** per CRI,s/p abx,management for sepsis,IVF,Afib control: s/p amio load,dc amio titrate metop to 50mg q8h,*****/steroids for CRS--> decadron per CRI,APAP for fevers,***** repletions,s/p vanc/mero (XX/XX/XX),Zosyn (XX/XX/XX) | ≤ 10 |

Table S16 Top 30 Clinical Intervention for Grade 2 ICANS

| **Clinical intervention** | **Frequency (patients)** |
| --- | --- |
| ***** | ≤ 10 |
| Received tocilizumab x 2 on XX/XX/XX and XX/XX/XX and dexamethasone 10 mg IV x 2 on XX/XX/XX. From XX/XX/XX-XX/XX/XX,his encephalopathy worsened and he was started on dexamethasone 10 mg IV Q6H,w/ subsequent significant improvement. | ≤ 10 |
| Tocilizumab 8 mg/kg (760 mg) x 1 on XX/XX/XX and XX/XX/XX - ***** 10 mg x 1 on XX/XX/XX then 10 mg Q6 (XX/XX/XX-XX/XX/XX) > 10 mg Q12 (XX/XX/XX-XX/XX/XX) > 10 mg/day (XX/XX/XX-XX/XX/XX) > sx resolved prior to discharge | ≤ 10 |
| Steroids treatment from XX/XX/XX to XX/XX/XX | ≤ 10 |
| Steroids XX/XX/XX-XX/XX/XX | ≤ 10 |
| Steroid treatment,Monitoring for symptoms,Thiamine administration,CT-guided LP | ≤ 10 |
| Started on steroids (dex 10mg IV BID),dex 10 mg q6h x 4 doses,dex 10 mg q8h x 3 doses,dex 10 mg q12h x 2 doses | ≤ 10 |
| Started on steroids (dex 10mg IV BID),Neuro checks,Taper plan: dex 10 mg q8h x 3 doses XX/XX/XX - dex 10 mg q12h x 2 doses XX/XX/XX - off dex after this,monitor for 24H for symptoms | ≤ 10 |
| Started on dexamethasone 10 mg IV Q6H | ≤ 10 |
| Started on dex 10 mg IV | ≤ 10 |
| Started on dex | ≤ 10 |
| Started on ***** 10mg IV Q6h on XX/XX/XX received a total of 3 doses then increased to 20mg IV Q6h x4 doses on XX/XX/XX | ≤ 10 |
| Started on ***** 10mg IV Q6h on XX/XX/XX received a total of 3 doses then increased to 20mg IV Q6h doses on XX/XX/XX. Solumedrol 1000mg x1 given XX/XX/XX,XX/XX/XX. Ordered to receive again on XX/XX/XX. Continue Anakinra 100mg SC daily (XX/XX/XX-) - ***** switched to Solumedrol 100mg IV q6hours on XX/XX/XX given concern that dex is contributing to severe bradycardia. | ≤ 10 |
| Started on ***** 10mg IV Q6h | ≤ 10 |
| Started dexamethasone x 4 doses,continued dex 10mg iv Q6hr (XX/XX/XX) | ≤ 10 |
| Started dexamethasone XX/XX/XX 10 mg IV q6 hrs | ≤ 10 |
| Started dex 10 q 12 x 6 doses | ≤ 10 |
| Started IV Dexamethasone 10 mg twice daily x 3 days | ≤ 10 |
| S/p Solumedrol 1gram IV daily x3 days (XX/XX/XX-XX/XX/XX) given highest c/f neurotoxicity,now on taper to 500mg IV on XX/XX/XX,and 250mg IV on XX/XX/XX,then off - Decadron 10mg q12h (XX/XX/XX - ) - S/p decadron 10mg q12h (XX/XX/XX - XX/XX/XX),s/p dex 10mg q6h (XX/XX/XX - XX/XX/XX) - Appreciate Neurology consultation - S/p Keppra load with 3grams IV x1 dose on XX/XX/XX,and increase standing Keppra to 1gram BID - OK to deescalate abx and defer LP - TTE,tele for stroke w/u negative - Delirium precautions - PT/OT/ST eval. PT eval'd XX/XX/XX w/ rec for home with PT | ≤ 10 |
| S/p ***** 10mg q6h (XX/XX/XX to XX/XX/XX,first dose at 0930 on XX/XX/XX),***** 10mg q12h (XX/XX/XX - 23),***** 10mg daily (XX/XX/XX - XX/XX/XX),Delirium precautions: open curtains during the day,***** ***** *****,minimize noise at night. | ≤ 10 |
| S/p ***** 10mg q6h (XX/XX/XX to XX/XX/XX,first dose at 0930 on XX/XX/XX),***** 10mg q12h (XX/XX/XX - 23),***** 10mg daily (XX/XX/XX - ),MRI Brain w wo contrast ordered XX/XX/XX (pending),EEG,Delirium precautions: open curtains during the day,***** ***** *****,minimize noise at night,Appreciate neuro consult | ≤ 10 |
| S/p ***** 10mg q6h (XX/XX/XX to XX/XX/XX,first dose at 0930 on XX/XX/XX) | ≤ 10 |
| Restarted dexamethasone 10mg IV BID | ≤ 10 |
| Restarted Dexamethasone 10mg IV BID,Neuro consultation,NCHCT and video EEG planned | ≤ 10 |
| Restarted Dexamethasone 10mg IV BID on XX/XX/XX and now with somewhat improving mental status,and concern for underlying CRS/neurotoxicity. Decrease Dexamethasone to 10mg IV q12 (XX/XX/XX--> ) (previously on XX/XX/XX 10 BID,XX/XX/XX 10q6,XX/XX/XX 10 TID). Keppra 500mg IV q12h (restart XX/XX/XX-) -- please keep on until at least day 30 (XX/XX/XX) | ≤ 10 |
| Restarted ***** 10mg daily x 3 days | ≤ 10 |
| Restarted ***** 10mg IV q8h (XX/XX/XX-,incr XX/XX/XX-,decr XX/XX/XX-),Restart keppra 500mg IV q12h (restart XX/XX/XX-) | ≤ 10 |
| Restarted ***** 10mg IV q6h (XX/XX/XX-,incr XX/XX/XX-),Restart keppra 500mg IV q12h (restart XX/XX/XX-) | ≤ 10 |
| Restarted ***** 10mg IV BID on XX/XX/XX,completed ***** taper on XX/XX/XX | ≤ 10 |
| Restarted ***** 10mg IV BID on XX/XX/XX and now with somewhat improving mental status,and concern for underlying CRS/neurotoxicity. Management: - Appreciate Neurology recs,now signed off - Completed ***** ***** on XX/XX/XX - Previously on 10q12 (XX/XX/XX-->XX/XX/XX ) XX/XX/XX 10 BID,XX/XX/XX 10q6,TID XX/XX/XX-XX/XX/XX then q12h XX/XX/XX-XX/XX/XX,10mg every day XX/XX/XX,then 4mg daily on XX/XX/XX then off) - Keppra 500mg IV q12h (restart XX/XX/XX-) -- please keep on until at least day 30 (XX/XX/XX) - See previous notes for ICE score history | ≤ 10 |

Table S17 All Clinical Intervention for Grade 2-3 ICANS

| **Clinical intervention** | **Frequency (patients)** |
| --- | --- |
| Dexamethasone 10 mg IV Q6H and ICU transfer | ≤ 10 |
| Dexamethasone 10 mg IV Q6H and ICU transfer for closer monitoring | ≤ 10 |
| Started on dexamethasone 10 mg IV Q6H for grade XX/XX/XX neurotoxicity,per the ***** FDA insert (at *****://*****.*****.*****/*****/*****.*****),Transferred to ICU for close monitoring | ≤ 10 |
| dexamethasone 10 mg IV Q6H and ICU transfer | ≤ 10 |
| dexamethasone 10 mg IV Q6H and transferred to the ICU | ≤ 10 |

Table S18 Top 30 Clinical Intervention for Grade 3 ICANS

| **Clinical intervention** | **Frequency (patients)** |
| --- | --- |
| 10mg decadron q6h | ≤ 10 |
| Started on ***** 20mg IV q6hours without improvement in symptoms. Neurology consulted,and felt EEG could be c/w seizures causing *****'s paralysis. He was loaded with Keppra. Steroid dosing increased to Solumedrol 1gram IV daily x3 days. | ≤ 10 |
| Solumedrol 1gram IV daily x3 days (XX/XX/XX-XX/XX/XX),now on taper to 500mg IV on XX/XX/XX,and 250mg IV on XX/XX/XX,then off | ≤ 10 |
| Start Dexamethasone 20mg IV Q6H | ≤ 10 |
| Start Solumedrol 1gram IV daily x3 days,Neurology consultation,ICU eval,transfer to ICU for closer monitoring and nursing care,Stat CTA brain and neck,Plan for MRI brain w/w/o contrast,Given Keppra load with 3grams IV x1 dose,and increase standing Keppra to 1gram BID,Start Meningitic dosing of abx with Acyclovir,Ceftriaxone,Vanc,Ampicillin,Plan for diagnostic LP in the AM after plts | ≤ 10 |
| Started on ***** 10mg IV Q6h on XX/XX/XX received a total of 3 doses then increased to 20mg IV Q6h doses on XX/XX/XX-> switched to Solumedrol 1000mg due to severe bradycardia XX/XX/XX dex (XX/XX/XX,XX/XX/XX,XX/XX/XX) - currently on Solumedrol taper over the next XX/XX/XX days decreasing the dose buy 50% every XX/XX/XX days. Started Taper on XX/XX/XX. Solumedrol 500mg IV daily x 2 days (XX/XX/XX-XX/XX/XX)-> solumedrol 125mg x1 on XX/XX/XX. - Continue Anakinra 100mg SC daily (XX/XX/XX-) | ≤ 10 |
| Started on ***** 10mg IV Q6h on XX/XX/XX received a total of 3 doses then increased to 20mg IV Q6h doses on XX/XX/XX. | ≤ 10 |
| Started on ***** 10mg IV Q6h on XX/XX/XX received a total of 3 doses then increased to 20mg IV Q6h doses on XX/XX/XX. - ***** switched to Solumedrol 100mg IV q6hours on XX/XX/XX given concern that dex is contributing to severe bradycardia. | ≤ 10 |
| Started on ***** 10mg IV Q6h on XX/XX/XX received a total of 3 doses then increased to 20mg IV Q6h doses on XX/XX/XX. Solumedrol 1000mg x1 given XX/XX/XX,XX/XX/XX. Ordered to receive again on XX/XX/XX. Continue Anakinra 100mg SC daily (XX/XX/XX-) - ***** switched to Solumedrol 100mg IV q6hours on XX/XX/XX given concern that dex is contributing to severe bradycardia. | ≤ 10 |
| Started on ***** 10mg IV Q6h on XX/XX/XX received a total of 3 doses then increased to 20mg IV Q6h doses on XX/XX/XX. Solumedrol 1000mg x1 given XX/XX/XX,XX/XX/XX. Ordered to receive again on XX/XX/XX. Now mental status has improved and is currently on Solumedrol taper over the next XX/XX/XX days decreasing the dose ***** 50% every XX/XX/XX days. Started Taper on XX/XX/XX. Solumedrol 500mg IV daily x 2 days (XX/XX/XX-XX/XX/XX) f/b 100mg IV daily etc. - Continue Anakinra 100mg SC daily (XX/XX/XX-) - ***** switched to Solumedrol 100mg IV q6hours on XX/XX/XX given concern | ≤ 10 |
| Started on ***** 10mg IV Q6h on XX/XX/XX received a total of 3 doses then increased to 20mg IV Q6h doses on XX/XX/XX. Solumedrol 1000mg x1 given XX/XX/XX. Plan to give Anakinra on XX/XX/XX - ***** switched to Solumedrol 100mg IV q6hours on XX/XX/XX given concern that dex is contributing to severe bradycardia. | ≤ 10 |
| Started on ***** 10mg IV Q6h on XX/XX/XX received a total of 3 doses then increased to 20mg IV Q6h x4 doses on XX/XX/XX | ≤ 10 |
| Started on ***** 20mg IV q6hours without improvement in symptoms. Neurology consulted,and felt EEG could be c/w seizures causing *****'s paralysis. | ≤ 10 |
| Started on Decadron 20mg IV q6h,Decrease Keppra back to 500mg PO BID,Follow up final blood cultures,Follow up MRI brain | ≤ 10 |
| Appreciate Neurology consultation | ≤ 10 |
| Started on Decadron 20mg IV q6h,ordered stat NCHCT,EEG,increased Keppra to 1g BID,and placed neurology consult for further eval | ≤ 10 |
| Treated with solumedrol pulse | ≤ 10 |
| Treated with solumedrol pulse,continued keppra | ≤ 10 |
| dex 10mg IV q6 hours (March - ),keppra BID D-1 through ***** | ≤ 10 |
| dexamethasone 20mg IV q6h (XX/XX/XX-XX/XX/XX) | ≤ 10 |
| methylpred 1g qday followed by taper XX/XX/XX - XX/XX/XX - MRI,EEG; if results inconclusive and not improving w/ steroids will consider LP - continue keppra BID - feeding tube placed XX/XX/XX for obtundation and nutritional support | ≤ 10 |
| s/p ***** 10mg IV x 1 on XX/XX/XX,and x3 on XX/XX/XX | ≤ 10 |
| s/p ***** x1 on XX/XX/XX ~1740,XX/XX/XX at 1530 | ≤ 10 |
| s/p solumedrol 1g IV daily x 3 doses (XX/XX/XX-XX/XX/XX) | ≤ 10 |
| s/p solumedrol 1g IV daily x 3 doses (XX/XX/XX-XX/XX/XX) then 250mg IV x 1 on XX/XX/XX | ≤ 10 |
| solumedrol 1g IV daily x 3 doses (XX/XX/XX-XX/XX/XX) then 250mg IV x 1 on XX/XX/XX | ≤ 10 |
| S/p Solumedrol 1gram IV daily x3 days (XX/XX/XX-XX/XX/XX) given highest c/f neurotoxicity,now on taper to 500mg IV on XX/XX/XX,and 250mg IV on XX/XX/XX,then off | ≤ 10 |
| S/p Keppra load with 3grams IV x1 dose on XX/XX/XX,and increase standing Keppra to 1gram BID | ≤ 10 |
| S/p ***** on XX/XX/XX and XX/XX/XX - S/p dex 10mg q6H (XX/XX/XX-XX/XX/XX) - S/p dex 10mg q12H (XX/XX/XX-XX/XX/XX) | ≤ 10 |
| Plan to bolus PRN for any soft pressures. | ≤ 10 |

Table S19 All Clinical Intervention for Grade 3-4 ICANS

| **Clinical intervention** | **Frequency (patients)** |
| --- | --- |
| LP today w/ TAPS,f/u gram stain,culture,cell count/diff,cytology,glucose,protein; continue keppra 500 BID; continue solumedrol ***** XX/XX/XX-XX/XX/XX,then taper; continue Tocilizumab ***** PRN for CRS; Abx as below | ≤ 10 |
| continue keppra 500 BID - continue solumedrol ***** XX/XX/XX-XX/XX/XX,then taper - continue Tocilizumab ***** PRN for CRS | ≤ 10 |
| continue keppra 500 BID - continue solumedrol ***** XX/XX/XX-XX/XX/XX,then taper - continue Tocilizumab ***** PRN for CRS - continue Abx as below | ≤ 10 |

Table S20 All Clinical Intervention for Grade 4 ICANS

| **Clinical intervention** | **Frequency (patients)** |
| --- | --- |
| Appreciate Neurology consultation | ≤ 10 |
| Continue Solumedrol 1gram IV daily x3 days (XX/XX/XX-XX/XX/XX) given highest c/f neurotoxicity,plan to taper to 500mg IV on XX/XX/XX,and 250mg IV on XX/XX/XX,then off | ≤ 10 |
| S/p Keppra load with 3grams IV x1 dose on XX/XX/XX,and increase standing Keppra to 1gram BID | ≤ 10 |
| S/p Solumedrol 1gram IV daily x3 days (XX/XX/XX-XX/XX/XX) given highest c/f neurotoxicity,now on taper to 500mg IV on XX/XX/XX,and 250mg IV on XX/XX/XX,then off | ≤ 10 |
| S/p Solumedrol 1gram IV daily x3 days (XX/XX/XX-XX/XX/XX) given highest c/f neurotoxicity,now on taper to 500mg IV on XX/XX/XX,and 250mg IV on XX/XX/XX,then off - Appreciate Neurology consultation - S/p Keppra load with 3grams IV x1 dose on XX/XX/XX,and increase standing Keppra to 1gram BID - OK to deescalate abx and defer LP - TTE,tele for stroke w/u negative - Delirium precautions - PT/OT/ST eval,need ARU placement. ***** send Covid tomorrow AM,with goal to discharge on XX/XX/XX. | ≤ 10 |
| Solumedrol 1gram IV daily x3 days (XX/XX/XX-XX/XX/XX),now on taper to 500mg IV on XX/XX/XX,and 250mg IV on XX/XX/XX,then off | ≤ 10 |
| Started on ***** 20mg IV q6hours without improvement in symptoms. Neurology consulted,and felt EEG could be c/w seizures causing *****'s paralysis. | ≤ 10 |
| Started on ***** 20mg IV q6hours without improvement in symptoms. Neurology consulted,and felt EEG could be c/w seizures causing *****'s paralysis. He was loaded with Keppra. Steroid dosing increased to Solumedrol 1gram IV daily x3 days. | ≤ 10 |
| Treatment with steroids (dex),tapered off by XX/XX/XX | ≤ 10 |
